# Machine Learning Analysis of Electronic Health Records Identifies Interstitial Lung Disease and Predicts Mortality in Patients with Systemic Sclerosis

**DOI:** 10.1101/2025.06.02.25328786

**Authors:** Alec K. Peltekian, Kevin M. Grudzinski, Bradford C. Bemiss, Jane E. Dematte, Carrie Richardson, Mary Carns, Kathleen Aren, Bashar Kadhim, Jean Paul Higeuro Sevilla, Changwan Ryu, Nikolay S. Markov, Natania S. Field, Mengou Zhu, Alexandra Soriano, Matthew Dapas, Harris Perlman, Aaron Gundersheimer, Kavitha C. Selvan, Ayush Kalia, Morgan Emokpae, Duncan F. Moore, Luke V. Rasmussen, John Varga, Krishnan Warrior, Catherine A. Gao, Richard G. Wunderink, G. R. Scott Budinger, Alok Choudhary, Alexander V. Misharin, Monique Hinchcliff, Ankit Agrawal, Anthony J. Esposito

**Affiliations:** Department of Computer Science, Northwestern University McCormick School of Engineering and Applied Science, Evanston, IL, United States; Division of Pulmonary and Critical Care, Northwestern University Feinberg School of Medicine, Chicago, IL, United States; Simpson Querrey Lung Institute for Translational Science, Northwestern University Feinberg School of Medicine, Chicago, IL, United States; Division of Rheumatology, Northwestern University Feinberg School of Medicine, Chicago, IL, United States; Department of Nephrology, Department of Medicine, Yale School of Medicine, New Haven, CT, United States; Clinical and Translational Research Accelerator, Department of Medicine, Yale School of Medicine, New Haven, CT, United States; Section of Pulmonary, Critical Care, and Sleep Medicine, Department of Medicine, Yale School of Medicine, New Haven, CT, United States; Department of Medicine, Northwestern University Feinberg School of Medicine, Chicago, IL, United States; Section of Rheumatology, Allergy & Immunology, Yale School of Medicine, New Haven, CT, United States; Department of Preventive Medicine, Northwestern University Feinberg School of Medicine, Chicago, IL, United States; Division of Rheumatology, University of Michigan Medical School, Ann Arbor, MI, United States; Department of Electrical and Computer Engineering, Northwestern University McCormick School of Engineering and Applied Science, Chicago, IL, United States

## Abstract

**Background:** Interstitial lung disease (ILD) affects *>*40% of patients with systemic sclerosis (SSc/scleroderma) and is the leading cause of disease-related mortality. Although therapies may slow progression, outcomes remain poor, partly because ILD is often detected after irreversible lung injury has occurred. Although chest computed tomography (CT) is a sensitive tool for ILD detection and is recommended at SSc diagnosis, it is oftentimes not performed and even less often performed serially. We sought to develop tools to predict ILD and mortality in patients with SSc using data routinely available in the electronic health record (EHR) to inform medical decision-making.

**Methods:** We analyzed longitudinal EHR data from two SSc cohorts: Northwestern University (1,169 participants; derivation cohort) and Yale University (376 participants; validation cohort). We identified clinical features from existing cohort-linked EHR queries composing a convenience sample of data from participants spanning decades rather than employing a single unified data collection effort. Three ILD experts independently reviewed CT reports and classified each as having or lacking ILD. To explore derivation cohort data structure, patients with *>*=3 forced vital capacity (FVC) results available were identified and stratified according to prevalent or absent ILD. Using unsupervised trajectory-based clustering exploratory analyses, we determined standardized patterns across groups. ML models were then developed using clinical EHR data as predictor variables and prevalent ILD and all-cause mortality as outcome variables. Model performance was assessed using area under the receiver operating characteristic curve (AUC).

**Results:** Seventy-four clinical features with low missingness, including demographic, vital sign, laboratory, and pulmonary function test data, were utilized for analyses. Four robust PFT trajectory clusters were identified that were associated with ILD prevalence and mortality in exploratory analyses. A ML model for ILD detection achieved an AUC of 0.832 and retained performance in the Yale cohort (AUC 0.754). In addition to established predictors such as autoantibodies and pulmonary function, the model identified routine laboratory measurements, including red cell distribution width (RDW), white blood cell count, and serum chloride, as important contributors. One-year mortality prediction achieved AUCs of 0.904 in the North-western cohort and 0.910 in the Yale cohort. Among patients with SSc-ILD, one-year mortality was predicted with AUCs of 0.744 and 0.902 in the Northwestern and Yale cohorts, respectively. Unexpectedly, we found that subtle laboratory abnormalities (such as change in RDW) contributed to predicting mortality.

**Conclusions:** Our prediction models comprised of widely available EHR data are useful tools to identify SSc patients at high risk for prevalent ILD and all-cause mortality. Integration of these models into clinical practice could enable scalable risk stratification and inform individualized ILD screening and monitoring strategies for SSc patients.

## Introduction

Systemic sclerosis (SSc) is a rare autoimmune disease that predominantly affects middle-aged adults characterized by progressive multi-organ fibrosis [1, 2]. Interstitial lung disease (ILD) is the most common pulmonary manifestation of SSc and the leading cause of death [3, 4]. Estimates of the prevalence of SSc-ILD range from 40–60% [5–8]. In most patients with SSc-ILD, lung injury is evident at the time of SSc diagnosis [9–11]; however, a significant proportion develop pulmonary manifestations later in the course of disease [12]. Furthermore, continuous incidence of new-onset ILD in patients with SSc has been described that remains stable for up to ten years from baseline [13]. Clinical practice guidelines conditionally recommend screening of patients with SSc at “increased risk” for ILD with pulmonary function testing (PFT) and chest computed tomography (CT) [14]. Nevertheless, they do not offer specific strategies to identify these high-risk individuals or for re-screening, leading to significant variation in practice. The optimal combination of risk factors for ILD prediction and estimates of risk for poor outcomes remains unknown. Data-driven strategies for risk stratification are therefore needed to improve detection and management of SSc-ILD.

The clinical course of SSc-ILD is vastly heterogeneous, although most will exhibit decline in lung function even years after diagnosis [15, 16]. Current clinical tools offer limited ability to stratify patients at highest risk for the development of SSc-ILD, progression of known disease, or adverse outcomes [17]. PFT and chest CT remain the cornerstones of diagnosis and monitoring, yet both modalities have important limitations—early ILD may be missed while lung function remains within normal ranges [18, 19]. Early initiation of treatment is important in modifying the disease course but is often guided by clinical gestalt rather than objective features [15, 20], resulting in difficult decisions regarding the timing of initiation or discontinuation of effective therapies [21–24]. As a result, SSc-ILD is frequently detected or treated after substantial, often irreversible lung injury has occurred [17]. These challenges are underscored by the finding that mortality from SSc-ILD has remained largely unchanged [25]. They also highlight a critical unmet need for personalized, scalable, data-driven approaches to disease management to improve early identification of SSc-ILD and stratify mortality risk to guide decision-making about treatment.

In this study, we developed ML models trained on multiple features from the EHR routinely collected in clinical practice for the care of SSc patients, irrespective of previously known associations with SSc-ILD. Our predictive models performed well in the dataset used for training and in an independent external validation cohort of SSc patients. They confirmed known risk factors for the development of and mortality from SSc-ILD, while also identifying novel biomarkers. Our findings suggest that these models may be useful in developing individualized screening, monitoring, and treatment plans for SSc patients.

## Methods

### Human Participants and Data Collection

This study was conducted in two observational, longitudinal patient registries from the Northwestern and Yale Scleroderma Programs. All human research was approved by the Northwestern University (STU00002669) and Yale University Institutional Review Boards (2000026608). All participants provided informed consent. Data for participants enrolled in the study were extracted from the Northwestern Electronic Data Warehouse, which is a continuously updated and searchable repository of EHR data [26]. Additional data were collected by study-associated research coordinators or physicians during patient encounters or extracted through patient chart review and captured in a study-specific REDCap database. To be included in either registry, participants had to fulfill either the 1980 American College of Rheumatology (ACR) or the 2013 ACR/European Alliance of Associations for Rheumatology criteria for SSc diagnosis [27, 28], depending on enrollment date. Disease onset was defined as the date of first non-Raynaud’s symptom attributable to SSc. Diagnoses of limited cutaneous SSc (lcSSc), diffuse cutaneous SSc (dcSSc), or systemic sclerosis sine scleroderma (SSS) were confirmed by adjudication of EHR data by board-certified rheumatologists (C.R., M.H.). Cases with mixed connective tissue disease or overlap with other connective tissue disease(s) were excluded. Mortality was determined by recorded dates of death or lung transplant from the EHR and REDCap database. A detailed explanation of each data source and its specific contribution to the dataset can be found in **Supplemental Section 1**.

### Data Pre-Processing and Feature Selection

We extracted clinical features such as age, sex, race, autoantibodies (anti-topoisomerase I [Scl-70], anticentromere [ACA], anti-RNA polymerase III [RNA Pol III]), pulmonary hypertension (PH) status (no PH, pre-capillary PH, post-capillary PH), and time-to-event intervals (e.g., time from SSc diagnosis to ILD diagnosis) from the EHR. A total of seventy-four features with sufficient data coverage were incorporated into ML modeling. These spanned demographics, vital signs, six-minute walk tests, PFTs, complete blood counts (CBC), autoantibodies, and chemistries (see **Supplemental Section 1**). Variables were derived from existing registry-linked EHR queries, representing a convenience sample of data compiled over past decades through various project-specific requests rather than through a single unified data collection effort. With the exception of established biomarkers such as SSc subtype, PFTs, and autoantibodies, feature inclusion was not adjudicated or supervised. Instead, the feature set reflected data routinely collected in the registry and was available at scale, which enabled the identification of unexpected associations with disease outcomes. Features were described by either continuous or categorical (binary or ordinal) data types.

Features were grouped into year-by-year aggregates based on the first measurement for each patient (**Supplemental Figure** S1). A participant’s first year would start at his/her first measurement and every following year was subsequently incremented. If multiple measurements occurred within the same year, values were averaged. Missingness was common due to the retrospective and heterogeneous nature of EHR data.

### Determination of ILD Diagnosis

Diagnoses of SSc-ILD were adjudicated by review of radiology reports of chest CT imaging by experts in ILD (A.J.E., B.C.B., J.E.D., J.P.H.S., Ch. R.) utilizing a three-reader method that has been previously described [27, 29]. Only reports, not CT images, were reviewed by the readers, as the aim of this study was to utilize EHR data alone for predictive modeling. In cases of disagreement between readers, consensus was obtained through collaborative discussion of the report. Readers were blinded to their initial assessment during consensus adjudication. Participants with persistent uncertainty of ILD diagnosis despite collaborative adjudication were labeled as “unknown” and excluded from subgroup analyses of SSc-ILD. Participants with CT reports composed the “CT subgroup” that was utilized for SSc-ILD analyses, and those without reports were excluded from this subgroup. Cases in which ILD was adjudicated to be present on the first available CT report were labeled as “prevalent ILD”. Cases in which ILD was adjudicated to be present on CT report after a prior CT without ILD were labeled as “incident ILD”.

### Trajectory-Based Clustering of FVC Patterns

To identify distinct longitudinal patterns of changes in lung function in SSc participants, we performed an unsupervised trajectory-based clustering analysis using longitudinal forced vital capacity (FVC) measurements. Participants with less than three FVC measurements were excluded to ensure reliable estimation of individual trajectories.

For each participant, we derived a set of summary trajectory features capturing both baseline lung function and longitudinal behavior, including baseline FVC (first available measurement), linear rate of FVC change over time (slope), mean FVC across follow-up, within-patient FVC variability (standard deviation), and total follow-up duration (years from first measurement). The rate of FVC change was estimated using ordinary least squares regression of FVC on time since first measurement. All trajectory features were standardized, and participants were clustered using k-means (k = 4) selected by expert adjudication. Cluster assignments were used to define distinct FVC trajectory patterns.

Associations between trajectory clusters and clinical outcomes were evaluated by comparing mortality rates and ILD prevalence across clusters. In addition to the full cohort analysis, we conducted a subgroup analysis restricted to participants with confirmed ILD. Within this ILD subgroup, the same clustering approach was applied, and cluster assignments were evaluated for associations with mortality and time to ILD diagnosis. Associations between trajectory clusters and categorical outcomes (mortality and ILD status) were assessed using chi-squared tests.

### Clustering and Phenotypic Analysis

Hierarchical clustering was applied to identify phenotypic groups using a subset of the EHR features (e.g., autoantibodies, SSc subtype, age at SSc diagnosis, and ILD status). We employed agglomerative hierarchical clustering using Euclidean distance metrics complete linkage method, implemented through the Morpheus package [30]. This approach maximizes the distance between elements of each cluster, resulting in more compact, well-separated phenotypic groupings. We determined that ten was the optimal number of clusters through iteratively increasing the number of clusters until the addition of a new cluster no longer provided clinically meaningful differentiation, as determined by a multidisciplinary team of SSc and ILD experts (A.J.E., J.E.D, B.C.B, C.R.). This approach enabled the identification of clusters of distinct participant attributes, which were subsequently analyzed for patterns related to development of ILD and mortality.

### Modeling Outcomes of Interest

All ML models were trained and internally validated using data from eligible participants from the Northwestern Scleroderma Registry (“Northwestern cohort”). The resultant models were subsequently deployed without retraining on clinical data from eligible participants in the Yale Rheumatology Program Patient Registry (“Yale cohort”) to assess external generalizability. The Yale cohort included participants fulfilling identical inclusion and exclusion criteria as the Northwestern cohort. Model performance in both cohorts was evaluated using identical preprocessing, feature sets, and evaluation metrics. The models were tasked with the following objectives:

#### Modeling Task 1 (ILD detection in participants with SSc)

A model was derived to identify whether a participant with SSc currently had ILD using solely annualized EHR data. The purpose of this task was to associate previously unrecognized or underutilized biomarkers from the EHR with disease.

#### Modeling Task 2 (mortality prediction in participants with SSc)

A model was derived to predict participant mortality within one, three, and five years from the first EHR data entry using annualized EHR data from all SSc participants. This model utilized ILD as a feature, which distinguishes it from Modeling Task 3.

#### Modeling Task 3 (mortality prediction in participants with known SSc-ILD)

A model was derived to predict mortality in a subgroup of participants with SSc-ILD within one, three, and five years after the adjudicated diagnosis using annualized EHR data. Data from up to one year prior to SSc-ILD diagnosis were included as baseline features in the mortality model. Subsequent follow-up data were aggregated into annual intervals after the date of SSc-ILD diagnosis (**Supplemental Figure** S1).

We applied the finalized models trained on the Northwestern cohort to participants enrolled in the Yale cohort for external validation.

### Statistical Methods

Comparative analyses between participant groups were conducted using the non-parametric Mann-Whitney U test for continuous variables given the potential for non-normally distributed data across features. Sample proportions were compared by chi-squared analysis. Kaplan-Meier survival curves were generated to assess time-to-event outcomes, and comparisons between groups were performed by the log-rank test. For predictive modeling tasks, model performance was evaluated using receiver operating characteristic (ROC) analysis with the area under the curve (AUC) reported to quantify discriminatory ability. All statistical tests were two-sided with significance defined at a threshold of p*<*0.05.

### Modeling and Framework

We employed Logistic Regression (LR) [31], Random Forest Classifier (RF) [32], XGBoost [33] LightGBM [34], and Neural Networks (NN) [35] machine learning (ML) algorithms to direct ILD detection and mortality prediction tasks. Hyperparameter optimization was conducted using Optuna, [36] with twenty-five trials per model-task pair. Model selection was based on five-fold cross-validation scores. Missing data were addressed with an imputation pipeline combining quartile binning (continuous features), mask-based encoding (missing features), and one-hot encoding (categorical features) [37] as described in **Supplemental Section 2**. Feature importance was evaluated using SHapley Additive exPlanations (SHAP) [38] and ablation studies, identifying key predictors and assessing robustness of the model to missing data. A detailed description of the model framework, imputation strategy, and optimization process can be found in **Supplemental Section 2**.

### Code and Data Availability Statement

The datasets generated and analyzed during this study are currently not publicly available; however, data are available for sharing upon reasonable request to the corresponding author and establishment of a data use agreement. The code used for data processing and model development is available on GitHub: https://github.com/NUPulmonary/SScILD-EHR-M1.

## Results

### Cohort Characteristics

#### All-Comers with SSc

We analyzed data from 1,169 SSc patients from the Northwestern Scleroderma Registry to serve as the derivation cohort for this study. Participants contributed EHR data encompassing a total of 15,494 person-years of observation with a median follow-up duration of 6.19 years. The Northwestern cohort was composed of predominantly White female participants with a median age of 45 years at the time of SSc diagnosis (Table 1). The majority of participants had lcSSc. Autoantibody analysis demonstrated positive Scl-70 in 27%, ACA in 23%, and RNA Pol III in 17%.

**Table 1:**
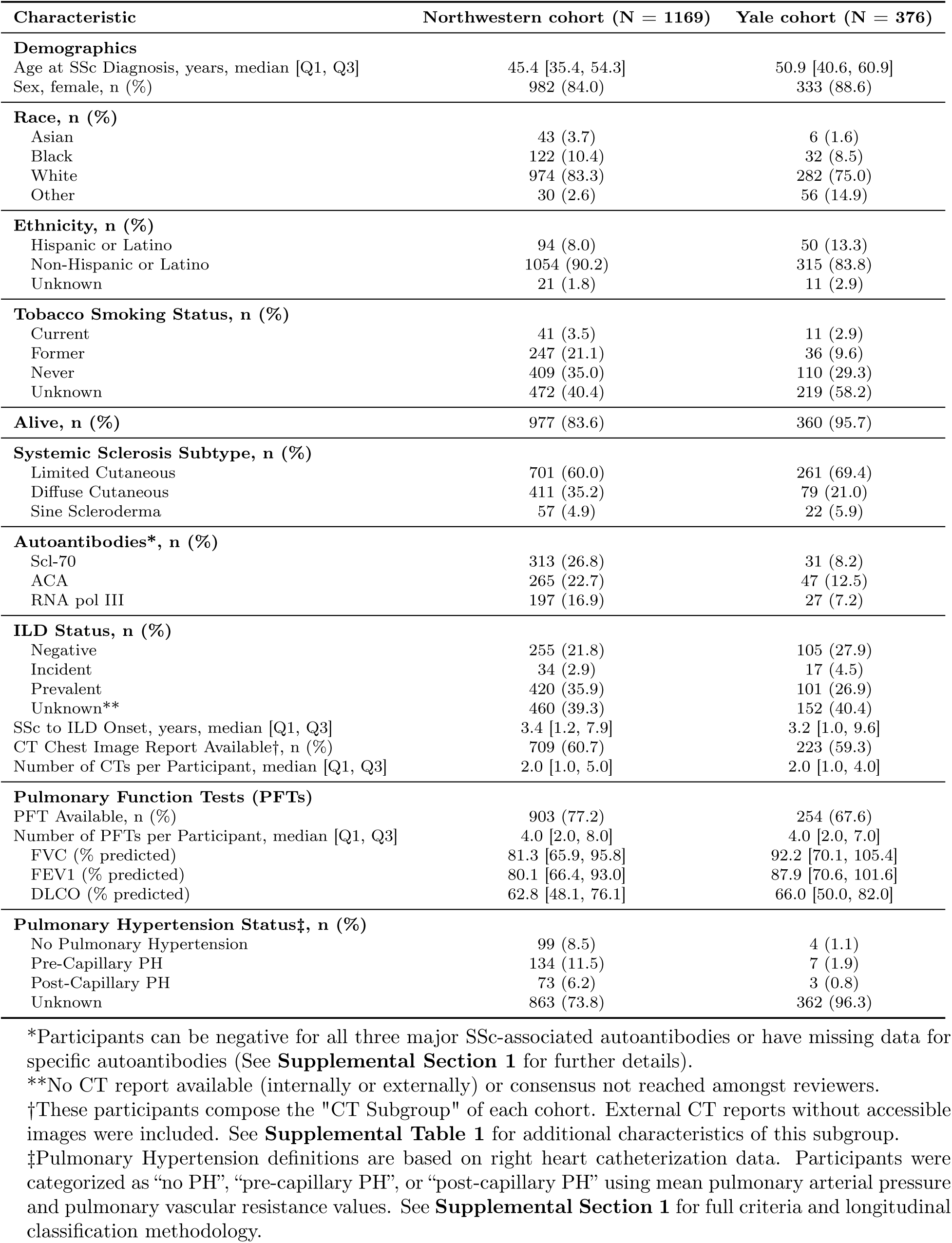
Cohort Characteristics.

Three hundred seventy-six participants with SSc comprised the Yale cohort, with a median follow-up duration of 6.17 years. 75.0% were White and 88.6% were female with a median age at SSc diagnosis of 51 years. Compared to the Northwestern cohort, the Yale cohort had a higher proportion of participants with lcSSc (69.4% vs. 59.9%) and a lower proportion with dcSSc (21.0% vs. 35.2%). Autoantibody prevalence differed as well, with lower proportions of Scl-70, ACA, and RNA Pol III. Similar to Northwestern, analyses of the Yale cohort revealed distinct autoantibody distributions, with lower Scl-70 positivity and comparable ACA positivity when restricting to patients with available serologic data (see Supplementary Table S1).

Participants in both cohorts were followed longitudinally for a median of 6.2 years. Detailed descriptions of cohort enrollment patterns, encounter frequency, and temporal growth of participation are provided in Supplemental Figure S2.

#### Participants with SSc-ILD

Of the 1169 participants in the Northwestern cohort, 709 (61%) had at least one CT (see Supplementary Table S1 for subgroup characteristics). The median number of chest CT reports available for review was 2 (interquartile range [IQR]: 1-4) per participant. Those with a CT were more likely to have dcSSc (42.7% vs. 35.2%) (p = 0.0041) and Scl-70 antibodies (p = 0.0013) compared to the total cohort. Of the 709 with CT reports, 454 (64%) were adjudicated as ILD (Supplemental Figure S3A). For 420 (93%), ILD was present on the first CT (i.e., prevalent ILD). Only 34 participants (7%) were diagnosed with ILD on a subsequent CT (i.e., incident ILD) [mean (standard deviation; SD) duration between negative baseline and positive follow-up CT for ILD was 4.07 years (4.52)]. Of the 376 Yale participants, 223 (59.3%) had at least one chest CT report available for review (median = 2, IQR: 1-4). Among those with chest CT reports (CT subgroup), 118 participants (53%) had an adjudicated diagnosis of ILD. For 101 participants (86%), ILD was present on the first CT, while 17 participants (14%) were diagnosed with ILD on a subsequent CT [mean (SD) duration between negative baseline and positive follow-up CT for ILD was 4.02 years (2.75)] (**Supplemental Figure** S3B, S4 - S7). Survival analyses of the CT subgroup of the Northwestern cohort demonstrated significantly reduced survival among participants with ILD (median 29.1 years) compared to those without ILD (median 36.9 years, log-rank p<0.0001; **Figure** 1).

**Figure 1:**
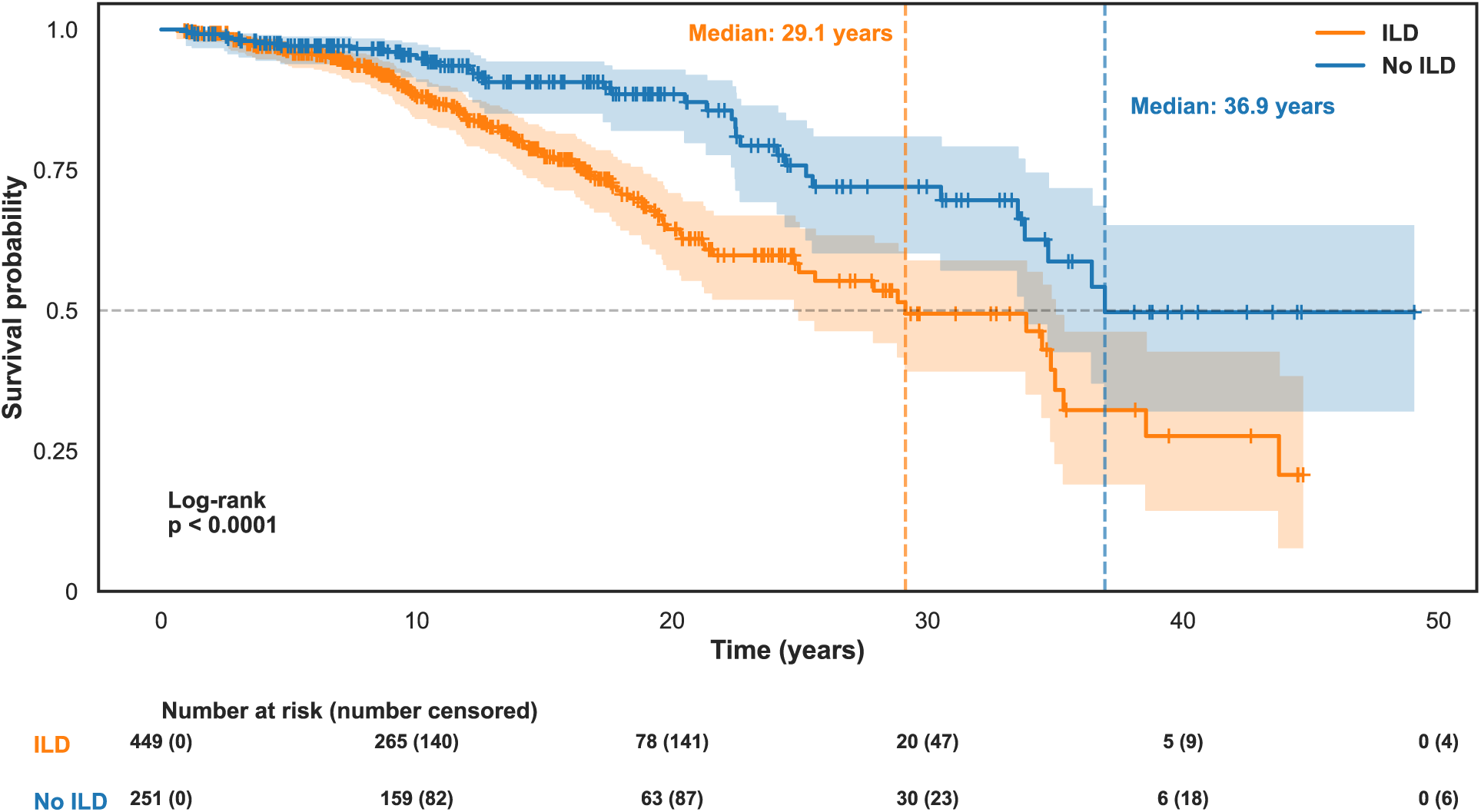
Kaplan-Meier survival curves for those with systemic sclerosis with or without interstitial lung disease (ILD) in the Northwestern cohort. Participants with ILD demonstrated significantly reduced survival compared to those without ILD. Shaded regions represent 95% confidence intervals.

### Cohort Phenotypic Clusters

To explore phenotypic heterogeneity among Northwestern cohort participants, we performed hierarchical clustering using disease-related features and other data including age at SSc diagnosis, autoantibodies, highest recorded modified Rodnan skin score (mRSS), and ILD diagnosis (**Supplemental Figure** S8A). Ten clusters were selected based on expert consensus of clinical interpretability and relevance (A.J.E., B.C.B., and J.E.D.).

In the Northwestern cohort, Cluster 6 was characterized by ACA, lcSSc, and absence of ILD — 88.3% of participants in this cluster were both ACA-positive and lcSSc. In contrast, participants with Scl-70 were largely concentrated in Clusters 7 and 8, which were dominated by dcSSc and higher prevalence of ILD (47.7% and 52.2%, respectively). Clusters 3 and 8 were enriched with participants with RNA Pol III who also demonstrated a dcSSc phenotype and high proportion of ILD (which agrees with recent reports [39, 40]). Similar clustering pattern was observed in the Yale cohort (**Supplemental Figure** S8B).

### FVC Trajectories and Clinical Outcomes

Unsupervised (k-means) clustering of longitudinal FVC measurements in the Northwestern cohort identified four distinct trajectory patterns distinguished by baseline lung function and rates of decline (Figure 2A–C). These trajectory clusters were associated with significantly different clinical outcomes. Mortality rates varied substantially across clusters, ranging from 8.5% to 43.5% (p*<*0.001). Clusters with lower baseline and more rapid decline in FVC demonstrated the highest burden of ILD and the poorest survival.

**Figure 2:**
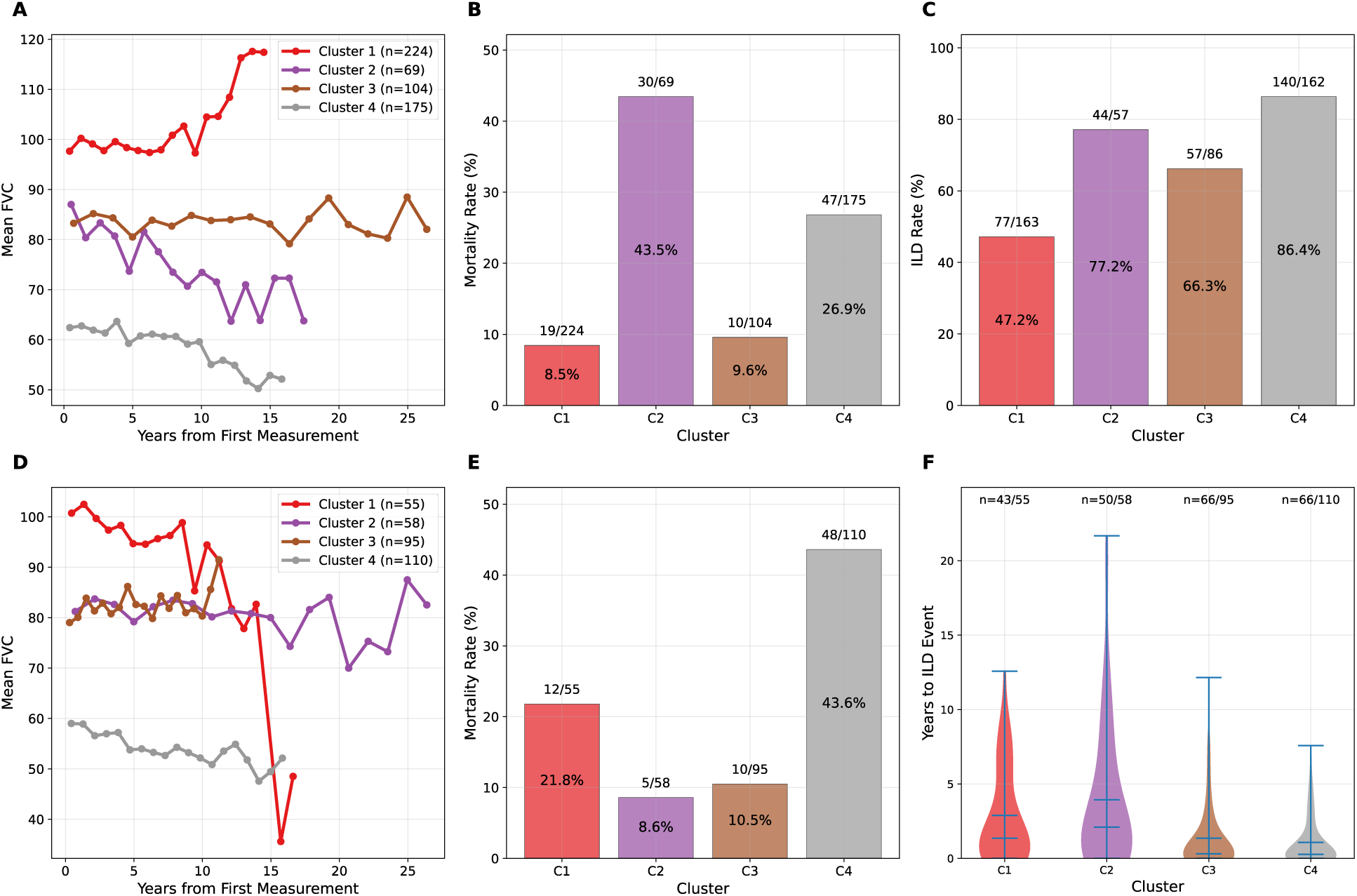
Trajectory-based clustering analysis by pulmonary function of participants with systemic sclerosis (SSc) and a subset with SSc-associated interstitial lung disease (ILD) reveals distinct clusters with differentially associated clinical outcomes in the Northwestern cohort. Unsupervised trajectory-based k-means clustering analysis of serial forced vital capacity (FVC) measurements (*n →* 3) and associated features (e.g., baseline measurement, slope, mean; see **Methods**) was conducted in the Northwestern cohort. Associations between clusters and clinical outcomes were assessed by chi-squared tests. (**A**) Four clusters demonstrated distinct longitudinal patterns of decline in FVC that were associated with (**B**) mortality and (**C**) ILD prevalence. (**D**) Subgroup clustering analysis of those with SSc-ILD revealed four cluster-specific FVC trajectories associated with (**E**) mortality and (**F**) time to ILD diagnosis.

We next repeated the trajectory-based clustering analysis restricted to participants with confirmed ILD in the Northwestern cohort (Figure 2D–F). Within this ILD subgroup, four trajectory clusters again emerged with distinct patterns of FVC decline. Mortality rates remained significantly different across clusters, ranging from 8.6% to 43.6% (p*<*0.001). In addition, time to ILD diagnosis varied across clusters, with earlier ILD onset observed among participants exhibiting more rapid declines in FVC. These exploratory findings motivated the use of ML models to integrate multidimensional EHR data beyond single-variable trajectories.

### ILD Detection

We developed ML models to detect ILD solely from EHR data using temporally standardized feature sets (Figure 3). Most features required minimal averaging, and among those that did, the coefficient of variation across yearly bins remained low (Supplemental Figure S9A–S9B). The LightGBM model achieved robust performance, with an AUC of 0.832 in the Northwestern cohort and 0.754 in the Yale cohort (Figure 3A). SHAP analysis highlighted autoantibodies, particularly ACA and Scl-70, as key detectors of ILD (e.g., ACA-negative participants were more likely to have ILD than those who were ACA-positive). Functional parameters (DLCO, FVC), PH, smoking status, and advanced age were also predictive of ILD (Figure 3B). External validation demonstrated consistent feature importance patterns with ACA, DLCO, and Scl-70 as well as laboratory markers such as white blood cell count and serum chloride retaining predictive importance (Figure 3C).

**Figure 3:**
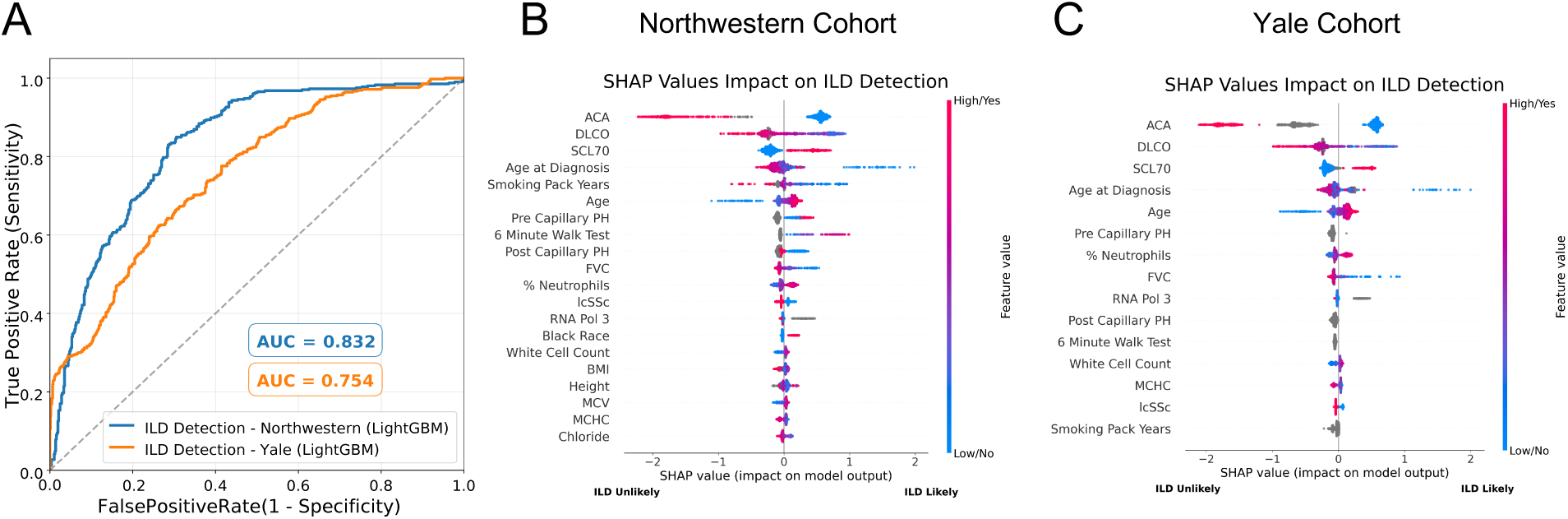
Model performance and feature importance for interstitial lung disease (ILD) detection in participants with systemic sclerosis. Machine learning algorithms were performed on features from the electronic health record. The LightGBM algorithm produced the best model performance. The model was derived in the Northwestern cohort and externally validated in the Yale cohort. (**A**) The receiver operating characteristic curves for model performance in both cohorts demonstrated strong discriminative ability. (**B**) SHapley Additive exPlanations feature importance analysis identifies those important for ILD detection in both the (**B**) Northwestern and (**C**) Yale cohorts.

### Mortality Prediction in Overall SSc Cohort

ML models were developed to predict one-, three-, and five-year mortality. A model using XGBoost with raw imputation (XGBoostRaw) achieved the best performance for the tasks of one-, three-, and five-year mortality prediction (AUC 0.904, 0.886, 0.857, respectively) in the Northwestern cohort (Figure 4A, Supplementary Figure S10A). The Yale cohort (Figure 4B) demonstrated strong generalizability of the resultant model with respective AUCs of 0.910, 0.920 and 0.867.

**Figure 4:**
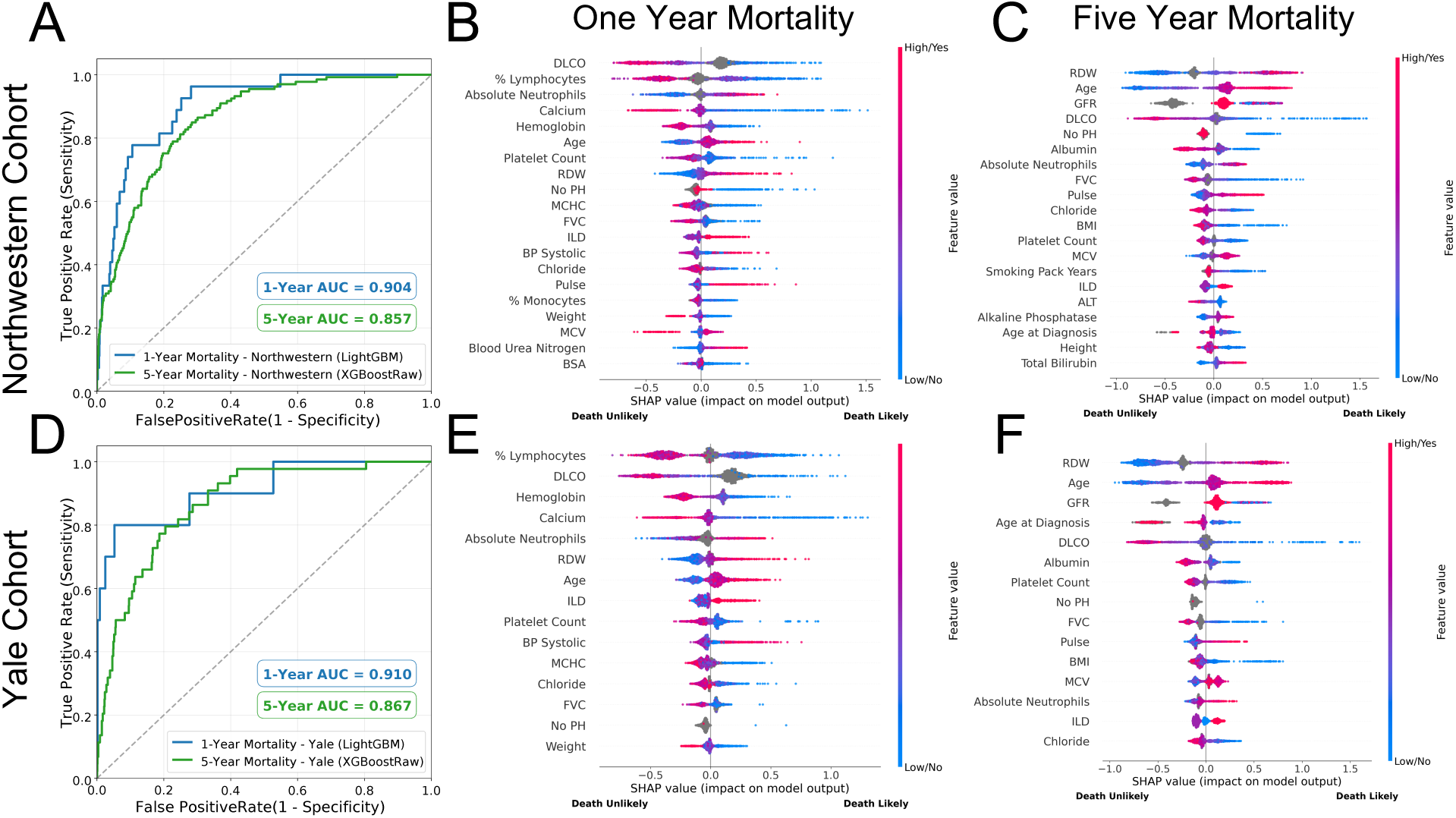
Model performance and feature importance for mortality prediction in participants with systemic sclerosis. Machine learning algorithms were performed on features from the electronic health record to predict mortality at one- and five-year timepoints (three-year prediction is displayed in Supplementary Figure S10A). The LightGBM algorithm produced the best model performance for one-year while the XGBoostRaw performed best for five-year performance. The models were derived in the Northwestern cohort and externally validated in the Yale cohort. (**A-B**) Receiver operating characteristic curves for mortality prediction over one- and five-year intervals demonstrated strong predictive performance that declined with longer time horizons. (**C-F**) SHapley Additive exPlanations feature importance plots highlight shifting importance of predictors from laboratory values in the short term (i.e., one-year mortality, [**C**, **E**] to chronic disease markers over longer observation (i.e., one-year mortality, [**D**, **F**] periods in both cohorts.

Feature importance analyses revealed dynamic shifts in predictors across time horizons. Clinical measurements classically associated with acute disease were the strongest drivers of one-year mortality risk (Figure 4C). These predictive features included hematologic indices (percent lymphocytes, absolute neutrophils, platelet count) and serum electrolyte levels (calcium). For three- and five-year mortality prediction models, chronic disease markers—including those associated with organ dysfunction (red cell distribution width [RDW], glomerular filtration rate [GFR]) and demographic factors (age)—were more important predictors (Figure 4D, Supplementary Figure S10C). PFT parameters (DLCO, FVC) remained important for predictive modeling across all time horizons. These features were similarly predictive in the Yale cohort (Figure 4E-F, Supplementary Figure S10D).

### Mortality Prediction in Participants with SSc-ILD

To explore the unique impact of ILD on mortality in SSc patients, we developed predictive ML models specific to participants with SSc-ILD. The XGBoostRaw model again demonstrated the highest performance for the tasks of one-, three, five-year mortality prediction in the Northwestern cohort (AUC 0.744, 0.785, 0.789) with similar performance in the Yale cohort (Figure 5, Supplementary Figure S10B).

**Figure 5:**
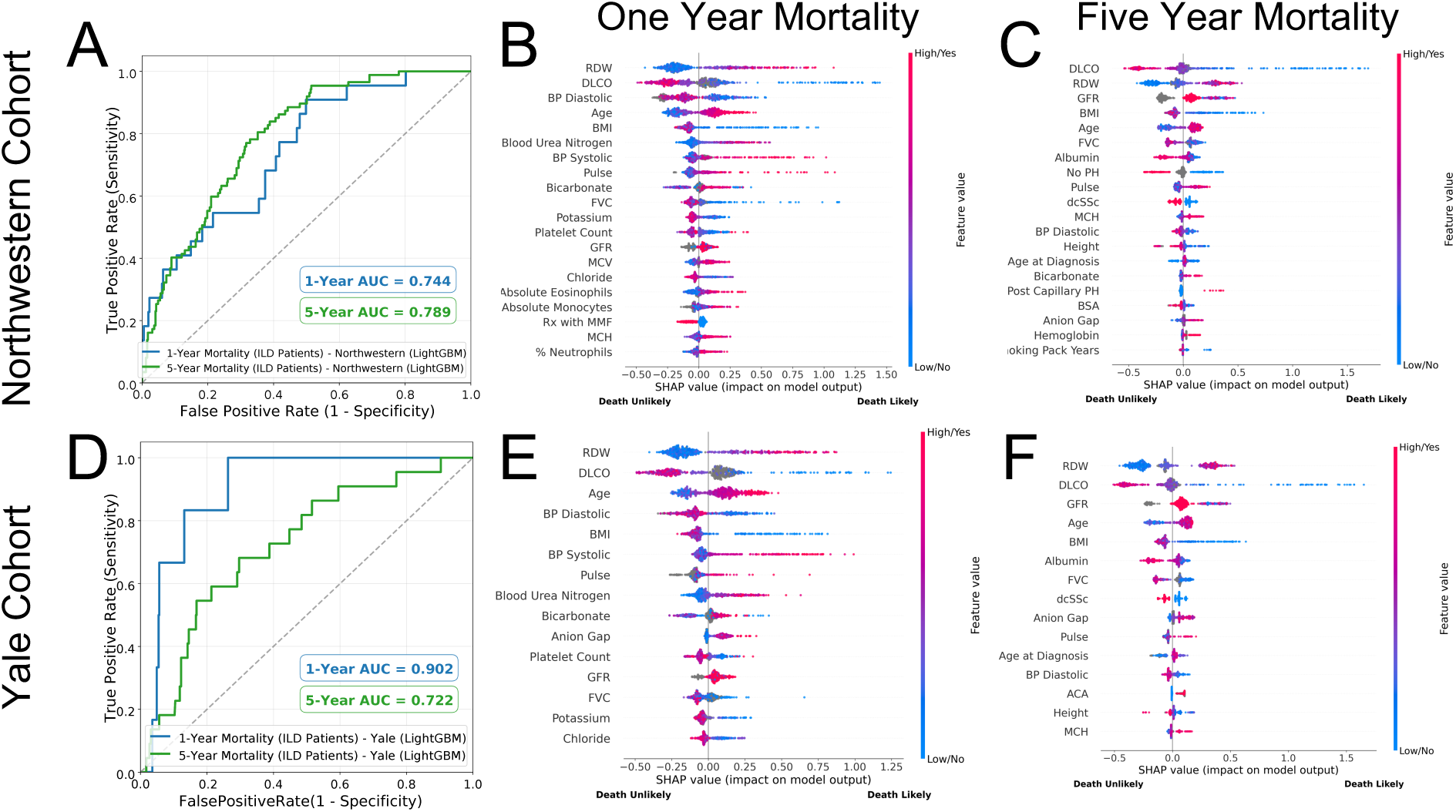
Model performance and feature importance for mortality prediction in participants with systemic sclerosis-associated interstitial lung disease (SSc-ILD). Machine learning algorithms were performed on features from the electronic health record to predict mortality at one- and five-year timepoints (three-year prediction is displayed in Supplementary Figure S10) for partipacipants with SSc-ILD. The LightGBM algorithm produced the best model performance for both one- and five-year predictive tasks. The models were derived in the Northwestern cohort and externally validated in the Yale cohort. (**A-B**)Receiver operating curves for mortality prediction in participants with confirmed SSc-ILD over one-and five-years demonstrated distinct patterns compared to mortality in the general SSc cohort (see Figure 4). (**C-F**) SHapley Additive exPlanations feature importance plots revealed evolving predictors, from vital signs and labs at one-year (**C**,**E**) to demographics and chronic disease markers by five-years (**D**, **F**) in both Northwestern (**C,D**) and Yale Cohorts (**E,F**).

Predictive models for short-term mortality in participants with SSc-ILD were similarly informed by acute markers of disease including vital signs and serum electrolyte levels (blood pressure (BP), pulse, bicarbonate, blood urea nitrogen), while those for long-term mortality were informed by chronic disease markers (albumin, RDW, GFR) and demographic factors (age, body mass index). Feature importance was consistent in the Yale cohort with PFT parameters (DLCO, FVC), organ dysfunction (GFR), and demographic factors again emerging as key predictors. Three-year mortality feature importance distributions are aligned with longer term predictions as displayed in Supplemental Figure S10E-F.

The resultant models were then tested on a combined dataset that contained both Northwestern and Yale cohorts. Model performance and key predictors remained stable across ILD detection and mortality prediction tasks (Supplemental Figures S11–S13). Features associated with both ILD diagnosis and mortality were often outside of the normal range (Supplemental Figure S14-S15A-E).

## Discussion

Our study leveraged large registries of participants with SSc at two tertiary care centers containing data collected longitudinally from the EHR. We employed ML algorithms to these data to detect ILD and to predict all-cause mortality in both all-comers with SSc and a subgroup of those with SSc-ILD. The resultant models yielded high predictive accuracy in assessing risk for these clinically important outcomes with AUCs ranging from 0.751 to 0.904. While the models identified important biomarkers previously associated with SSc-ILD progression such as impaired pulmonary function and autoantibody status, many were unexpected or under-recognized (e.g., RDW, GFR, serum chloride/calcium levels). While these measurements are routinely collected in the care of patients with SSc, they have predictive value at levels that may fall within the normal range and hence might be overlooked in a busy clinical practice. The ability of our models to highlight these biomarkers underscores the potential of ML to refine risk stratification and ultimately guide early interventions for SSc-ILD.

Our results demonstrate that most cases of ILD (86.0-92.5%) were detected on a participant’s first available CT. Incident ILD during observation was observed in only 4.8-7.7% of the cohorts. These results align with prior findings from the EUSTAR Group, which reported that only 8.2% of participants developed ILD during follow-up [41]. This underscores the importance of recently published ACR/American College of Chest Physicians Screening Guidelines conditionally recommending CT screening for ILD at the time of diagnosis of SSc [14]; however, it also demonstrates a need to stratify patients at-risk for incident ILD who would benefit from re-screening after an initial negative screening CT. Our models’ findings have the potential to guide clinicians in who would be best suited for re-screening based on clinical biomarkers in the EHR so that diagnosis of incident ILD is not delayed.

Current diagnosis and monitoring of SSc-ILD is limited primarily to PFTs and CT scans of the chest, the latter of which remains the gold standard. Guidelines from professional societies conditionally recommend baseline screening with both modalities at diagnosis of SSc patients at “high risk” for ILD; however, they do not provide explicit guidance on the optimal combination of risk factors that indicate high risk or when to repeat imaging in patients with negative initial studies to screen for incident ILD [14], which develops in a significant proportion of patients [12]. A similar dilemma surrounds timing and selection of therapies for SSc-ILD, many of which have demonstrated efficacy in altering the course of disease [21–23]. While there is mounting evidence that most patients with SSc-ILD will progress, [15, 16] clinical tools to identify those at high-risk are lacking.

We performed trajectory-based clustering analysis that highlighted the heterogeneous nature of lung function decline in SSc. Individual clusters may serve as prognostic indicators, particularly in identifying patients at risk for early-onset ILD; however they rely on features of a single variable. To capture non-linear interactions between multiple variables, we employed ML algorithms to data readily available in the EHR to create models that detect ILD and predict all-cause mortality in both all-comers with SSc and a subgroup of those with SSc-ILD. ML is capable of integrating large volumes of data and may aid clinicians in identifying those who are high risk and guide personalized clinical decision-making [42–44]. Conversely, ML may also identify those at low risk for poor clinical outcomes and provide the clinician with guidance on which patients may be appropriate for discontinuation of immunomodulatory therapy, a common scenario that currently has little guidance in clinical practice outside expert opinion [20].

Unsurprisingly, established predictors of SSc-ILD—including negative ACA, positive Scl-70, and lower DLCO—were among the most important features in our ML models for ILD detection. However, our analyses also identified routine clinical measurements, such as serum chloride level, white blood cell count, and RDW, as predictive of ILD. These clinical measurements—readily available in the EHR—may be commonly overlooked during evaluation of ILD, as they are not classic markers of disease and values are often in the normal range. In previous studies, investigators identified an association between RDW and changes in FVC and DLCO in participants with SSc-ILD and other connective tissue diseases [45, 46]. Our findings provide an unbiased validation of these observations and suggest that commonly measured EHR features could provide additional diagnostic and prognostic value and possibly insights into disease pathogenesis.

In assessing mortality risk, our models demonstrated high predictive performance, particularly for one-year mortality (AUC 0.904 in both cohorts). In short-term predictions (e.g., one-year mortality), laboratory values associated with acute disease—such as percent lymphocyte differential, absolute neutrophil count, serum calcium concentration, and serum chloride concentration—were the strongest markers of risk, suggesting that rapidly deteriorating clinical states contribute significantly to early mortality [47]. However, in longer-term prediction models (three- and five-year mortality models), chronic disease markers—including DLCO, RDW, age, and GFR—were more prominent, suggesting that persistent physiologic decline and demographic factors are important determinants of long-term survival.

When assessing mortality risk specifically among SSc-ILD participants, our models exhibited moderate predictive accuracy (AUC 0.744-0.789 and AUC 0.798-0.919 in Northwestern and Yale Cohorts, respectively), with vital signs and laboratory markers predominating in early mortality prediction and PFT parameters mounting importance in later outcomes. These data suggest that while acute clinical events may precipitate early mortality in ILD patients, long-term survival is dictated by chronic respiratory decline. The strong association between DLCO and mortality risk reinforces a loss of capillary surface area available for gas exchange as a key predictor of outcomes in ILD, [48] further supporting the use of DLCO in the management of SSc-ILD.

Our study has several limitations. First, model derivation was performed in a retrospective single-center cohort, which may introduce referral and selection bias particularly in terms of ILD prevalence and follow-up patterns. To address these concerns, we performed external validation using an independent cohort, which confirmed robust model performance across all tasks. Importantly, feature importance patterns remained consistent across cohorts, supporting the reliability of identified predictors. These validation results strengthen confidence in the broader applicability of our ML approach to diverse SSc populations. Second, ILD classification was based on expert adjudication of CT reports rather than direct imaging analysis, which could introduce inter-reader variability. This methodology was implemented intentionally, as the goal of our study was to utilize available EHR data and not rely on physician assessment of raw images. The methods employed, therefore, could lead to classification bias of ILD. However, we believe that use of only the EHR data allows this model to be translated into settings that lack expert adjudication of ILD diagnoses, expanding its use beyond tertiary care centers. Next, the sporadic and retrospective nature of EHR data presents a challenge in ML modeling. For example, PFT, CT, and laboratory data were collected at varying intervals and not at random. Our model approached this unstructured dataframe by averaging values annually, which may have led to insensitive attention to variation in data over a short timeframe. Regardless, our models were able to robustly predict patient outcomes despite the averaging we employed and the coefficient of variation of the features was low. Future work should explore approaches that avoid data averaging (for example, transformer-based methods) and the integration of deep learning-based image analysis to enhance diagnostic accuracy [49, 50]. Finally, our models relied on structured EHR data downloaded from a repository, which may underrepresent important clinical features that are captured in unstructured physician notes, such as patient-reported symptoms or nuanced disease severity assessments. Additional studies should consider use of natural language processing approaches to extract these data that may further refine predictive performance of our models.

## Conclusion

In this multicenter study, we demonstrate that ML models applied to routinely collected electronic health record data can accurately identify SSc-ILD and predict all-cause mortality across independent cohorts. By integrating longitudinal clinical, laboratory, and physiologic features, these models capture both established risk factors and additional markers of disease severity that are not routinely incorporated into clinical risk assessment. Although prospective validation is required, our findings suggest that EHR-based ML approaches could support risk stratification, inform individualized surveillance and proactive management strategies, and complement existing guideline-recommended tools in the care of patients with SSc.

## Data Availability

All data produced and analyzed in the present work are currently not publicly available due to participant privacy. The corresponding author will consider reasonable requests for data on an individual basis. The code used for data processing and model development is available on GitHub.

https://github.com/NUPulmonary/SScILD-EHR-M1

## Acknowledgements

This research was supported in part through a generous gift from K. Querrey and L. Simpson. This research was also supported by the computational resources and staff contributions provided for the Quest high-performance computing facility at Northwestern University, which is jointly supported by the Office of the Provost, the Office for Research, and Northwestern University Information Technology. This research was also supported in part through the computational resources and staff contributions provided by the Genomics Compute Cluster, which is jointly supported by the Feinberg School of Medicine, the Center for Genetic Medicine, Feinberg’s Department of Biochemistry and Molecular Genetics, the Office of the Provost, the Office for Research, and North-western Information Technology. The Genomics Compute Cluster is part of Quest, Northwestern University’s high-performance computing facility, with the purpose of advancing research in genomics. This work is dedicated to the benefit of all sentient beings. Ch.R. is supported by the NIH/NHLBI (grant no. K08HL151970), and the Boehringer Ingelheim Discovery Award. N.S.M. is supported by the American Heart Association (grant no. 24PRE1196998). L.V.R. is supported by NIH/NIAID U19 AI135964. CAG is supported by NIH/NHLBI K23HL169815, a Parker B. Francis Opportunity Award, and an American Thoracic Society Unrestricted Grant. R.G.W. is supported by NIH/NIAID U19 AI135964, RO1 AI158530; NHLBI RO1 HL149883, P01 HL154998; U01TR003528. G.R.S.B. is supported by the NIH (U19AI135964, P01AG049665, R01HL147575, P01HL071643, and R01HL154686); the US Department of Veterans Affairs (I01CX001777); a grant from the Chicago Biomedical Consortium; and a Northwestern University Dixon Translational Science Award. A.C. is supported by NSF (grant no. OAC-2331329). A.V.M. is supported by the NIH (grant nos. U19AI135964, P01AG049665, P01HL154998, U19AI181102, R01HL153312, R01HL158139, R01ES034350 and R21AG075423) and research grants from AbbVie and Merck. M.H. is supported by the NIH/NHLBI (grant no. R01AR07327). A.A. is supported by NIH grants (U19AI135964 and R01HL158139), SQLIFTS, and NSF grant OAC-2331329, and a grant from Pulmonary Fibrosis Foundation. A.J.E. is supported by the NIH (grant no. L30HL149048) and a grant from the Pulmonary Fibrosis Foundation. The funders had no role in the study design, data collection and analysis, decision to publish, or preparation of the manuscript.

## CONTRIBUTIONS

A.P., A.V.M. and A.J.E. conceived and designed the study. A.P. K.M.G., B.C.B., J.E.D., C.R., M.C., K.A., N.S.F., M.Z., A.S., M.D., A.G., K.S., D.M., J.V., M.E.H., and K.W. collected and curated data. A.P., B.C.B., J.E.D, G.R.S.B., A.V.M., A.A., and A.J.E. analyzed and interpreted the data. A.P., C.A.G., R.G.W., A.C. contributed to development of analytical approaches. A.P., B.K., J.P.H.S., M.E., A.V.M., and M.H. contributed to external validation processing and collection. H.P., M.E.H., and A.V.M. provided funding for the study. A.P., A.V.M., A.A., and A.J.E. wrote the manuscript. All authors read, edited, and approved the final manuscript.

## References

[1] Elizabeth R Volkmann, Kristofer Andréasson, and Vanessa Smith. Systemic sclerosis. The Lancet, 401(10373):304–318, 2023.

[2] Dinesh Khanna, Donald P Tashkin, Christopher P Denton, Elisabetta A Renzoni, Sujal R Desai, and John Varga. Etiology, risk factors, and biomarkers in systemic sclerosis with interstitial lung disease. American journal of respiratory and critical care medicine, 201(6):650–660, 2020.

[3] Muriel Elhai, Christophe Meune, Marouane Boubaya, Jérôme Avouac, Eric Hachulla, Alexandra Balbir-Gurman, Gabriela Riemekasten, Paolo Airò, Beatriz Joven, Serena Vettori, et al. Mapping and predicting mortality from systemic sclerosis. Annals of the rheumatic diseases, 76(11):1897–1905, 2017.

[4] Anthony J Tyndall, Bettina Bannert, Madelon Vonk, Paolo Airò, Franco Cozzi, Patricia E Carreira, Dominique Farge Bancel, Yannick Allanore, Ulf Müller-Ladner, Oliver Distler, et al. Causes and risk factors for death in systemic sclerosis: a study from the eular scleroderma trials and research (eustar) database. Annals of the rheumatic diseases, 69(10):1809–1815, 2010.

[5] Walker Ua. Clinical risk assessment of organ manifestations in systemic sclerosis: a report from the eular scleroderma trials and resesrch group database. Ann Rheum Dis, 66(6):754–763, 2007.

[6] Anna-Maria Hoffmann-Vold, Håvard Fretheim, Anne-Kristine Halse, Marit Seip, Helle Bitter, Marianne Wallenius, Torhild Garen, Anne Salberg, Cathrine Brunborg, Øyvind Midtvedt, et al. Tracking impact of interstitial lung disease in systemic sclerosis in a complete nationwide cohort. American journal of respiratory and critical care medicine, 200(10):1258–1266, 2019.

[7] Aurore Bergamasco, Nadine Hartmann, Laura Wallace, and Patrice Verpillat. Epidemiology of systemic sclerosis and systemic sclerosis-associated interstitial lung disease. Clinical epidemiology, pages 257–273, 2019.

[8] Rikisha Shah Gupta, Ardita Koteci, Ann Morgan, Peter M George, and Jennifer K Quint. Incidence and prevalence of interstitial lung diseases worldwide: a systematic literature review. BMJ Open Respiratory Research, 10(1), 2023.

[9] Veronika K Jaeger, Elina G Wirz, Yannick Allanore, Philipp Rossbach, Gabriela Riemekasten, Eric Hachulla, Oliver Distler, Paolo Airò, Patricia E Carreira, Alexandra Balbir Gurman, et al. Incidences and risk factors of organ manifestations in the early course of systemic sclerosis: a longitudinal eustar study. PloS one, 11(10):e0163894, 2016.

[10] Anna-Maria Hoffmann-Vold, Trond M Aaløkken, May Brit Lund, Torhild Garen, Øyvind Midtvedt, Cathrine Brunborg, Jan Tore Gran, and Øyvind Molberg. Predictive value of serial high-resolution computed tomography analyses and concurrent lung function tests in systemic sclerosis. Arthritis & rheumatology, 67(8):2205–2212, 2015.

[11] Oliver Distler, Shervin Assassi, Vincent Cottin, Maurizio Cutolo, Sonye K Danoff, Christopher P Denton, Jörg HW Distler, Anna-Maria Hoffmann-Vold, Sindhu R Johnson, Ulf Müller Ladner, et al. Predictors of progression in systemic sclerosis patients with interstitial lung disease. European Respiratory Journal, 55(5), 2020.

[12] Sabrina Hoa, Claudie Berger, Nouha Lahmek, Maggie Larché, Mohammed Osman, May Choi, Janet Pope, Carter Thorne, Canadian Scleroderma Research Group, M Baron, et al. Characterization of incident interstitial lung disease in late systemic sclerosis. Arthritis & Rheumatology, 77(4):450–457, 2025.

[13] Liubov Petelytska, Lorenzo Tofani, Arthiha Velauthapillai, Rucsandra Dobrota, Mike Oliver Becker, Carina Mihai, Sinziana Muraru, Muriel Elhai, Suzana Jordan, Eric Hachulla, et al. The incidence of interstitial lung disease in patients with systemic sclerosis: rate, risk factors and prognostic implications in a eustar cohort analysis (cp 133). Annals of the Rheumatic Diseases, 2026.

[14] Sindhu R Johnson, Elana J Bernstein, Marcy B Bolster, Jonathan H Chung, Sonye K Danoff, Michael D George, Dinesh Khanna, Gordon Guyatt, Reza D Mirza, Rohit Aggarwal, et al. 2023 american college of rheumatology (acr)/american college of chest physicians (chest) guideline for the treatment of interstitial lung disease in people with systemic autoimmune rheumatic diseases. Arthritis & rheumatology, 76(8):1182–1200, 2024.

[15] Moritz Scheidegger, Marouane Boubaya, Alexandru Garaiman, Imon Barua, Mike Becker, Hilde Jenssen Bjørkekjær, Cosimo Bruni, Rucsandra Dobrota, Håvard Fretheim, Suzana Jordan, et al. Characteristics and disease course of untreated patients with interstitial lung disease associated with systemic sclerosis in a real-life two-centre cohort. RMD open, 10(1):e003658, 2024.

[16] Anna-Maria Hoffmann-Vold, Yannick Allanore, Margarida Alves, Cathrine Brunborg, Paolo Airó, Lidia P Ananieva, László Czirják, Serena Guiducci, Eric Hachulla, Mengtao Li, et al. Progressive interstitial lung disease in patients with systemic sclerosis-associated interstitial lung disease in the eustar database. Annals of the rheumatic diseases, 80(2):219–227, 2021.

[17] Surabhi Agarwal Khanna, John W Nance, and Sally A Suliman. Detection and monitoring of interstitial lung disease in patients with systemic sclerosis. Current Rheumatology Reports, 24 (5):166–173, 2022.

[18] Elana J Bernstein, Sara Jaafar, Shervin Assassi, Robyn T Domsic, Tracy M Frech, Jessica K Gordon, Rachel J Broderick, Faye N Hant, Monique E Hinchcliff, Ami A Shah, et al. Performance characteristics of pulmonary function tests for the detection of interstitial lung disease in adults with early diffuse cutaneous systemic sclerosis. Arthritis & rheumatology, 72(11): 1892–1896, 2020.

[19] Kimberly Showalter, Aileen Hoffmann, Gerald Rouleau, David Aaby, Jungwha Lee, Carrie Richardson, Jane Dematte, Rishi Agrawal, Rowland W Chang, and Monique Hinchcliff. Performance of forced vital capacity and lung diffusion cutpoints for associated radiographic interstitial lung disease in systemic sclerosis. The Journal of rheumatology, 45(11):1572–1576, 2018.

[20] Franck F Rahaghi, Vivien M Hsu, Robert J Kaner, Maureen D Mayes, Ivan O Rosas, Rajan Saggar, Virginia D Steen, Mary E Strek, Elana J Bernstein, Nitin Bhatt, et al. Expert consensus on the management of systemic sclerosis-associated interstitial lung disease. Respiratory research, 24(1):6, 2023.

[21] Oliver Distler, Kristin B Highland, Martina Gahlemann, Arata Azuma, Aryeh Fischer, Maureen D Mayes, Ganesh Raghu, Wiebke Sauter, Mannaig Girard, Margarida Alves, et al. Nintedanib for systemic sclerosis–associated interstitial lung disease. New England Journal of Medicine, 380(26):2518–2528, 2019.

[22] Donald P Tashkin, Michael D Roth, Philip J Clements, Daniel E Furst, Dinesh Khanna, Eric C Kleerup, Jonathan Goldin, Edgar Arriola, Elizabeth R Volkmann, Suzanne Kafaja, et al. Mycophenolate mofetil versus oral cyclophosphamide in scleroderma-related interstitial lung disease (sls ii): a randomised controlled, double-blind, parallel group trial. The Lancet Respiratory Medicine, 4(9):708–719, 2016.

[23] Dinesh Khanna, Celia JF Lin, Daniel E Furst, Jonathan Goldin, Grace Kim, Masataka Kuwana, Yannick Allanore, Marco Matucci-Cerinic, Oliver Distler, Yoshihito Shima, et al. Tocilizumab in systemic sclerosis: a randomised, double-blind, placebo-controlled, phase 3 trial. The Lancet Respiratory Medicine, 8(10):963–974, 2020.

[24] Toby M Maher, Shervin Assassi, Arata Azuma, Vincent Cottin, Anna-Maria Hoffmann-Vold, Michael Kreuter, Justin M Oldham, Luca Richeldi, Claudia Valenzuela, Marlies S Wijsenbeek, et al. Nerandomilast in patients with progressive pulmonary fibrosis. New England Journal of Medicine, 392(22):2203–2214, 2025.

[25] Elizabeth R Volkmann, Donald P Tashkin, Myung Sim, Ning Li, Ellen Goldmuntz, Lynette Keyes-Elstein, Ashley Pinckney, Daniel E Furst, Philip J Clements, Dinesh Khanna, et al. Short-term progression of interstitial lung disease in systemic sclerosis predicts long-term survival in two independent clinical trial cohorts. Annals of the rheumatic diseases, 78(1):122–130, 2019.

[26] Ahmed Allam, Aron N Horvath, Matthias Dittberner, Cécile Trottet, Elise Siegert, Vincent Sobanski, Patricia Carreira Delgado, Dagna Lorenzo, Vanessa Smith, Serena Guiducci, et al. Predicting interstitial lung disease progression in patients with systemic sclerosis using attentive neural processes-a eustar study. medRxiv, pages 2024–04, 2024.

[27] Against Rheumatism Collaborative Initiative. 2013 classification criteria for systemic sclerosis. Arthritis & Rheumatism, 65(11):2737–2747, 2013.

[28] Alfonse T Masi, Subcommittee For Scleroderma Criteria of the American Rheumatism Association Diagnostic, and Therapeutic Criteria Committee. Preliminary criteria for the classification of systemic sclerosis (scleroderma). Arthritis & Rheumatism, 23(5):581–590, 1980.

[29] George R Washko, Gary M Hunninghake, Isis E Fernandez, Mizuki Nishino, Yuka Okajima, Tsuneo Yamashiro, James C Ross, Raúl San José Estépar, David A Lynch, John M Brehm, et al. Lung volumes and emphysema in smokers with interstitial lung abnormalities. New England Journal of Medicine, 364(10):897–906, 2011.

[30] Broad Institute. Morpheus. https://software.broadinstitute.org/morpheus, 2025. Accessed: 2025-10-09.

[31] David W Hosmer, Stanley Lemeshow, and Rodney X Sturdivant. Introduction to the logistic regression model. Applied logistic regression, 2:1–30, 2000.

[32] Leo Breiman. Random forests. Machine learning, 45(1):5–32, 2001.

[33] Tianqi Chen and Carlos Guestrin. Xgboost: A scalable tree boosting system. In Proceedings of the 22nd acm sigkdd international conference on knowledge discovery and data mining, pages 785–794, 2016.

[34] Guolin Ke, Qi Meng, Thomas Finley, Taifeng Wang, Wei Chen, Weidong Ma, Qiwei Ye, and Tie-Yan Liu. Lightgbm: A highly efficient gradient boosting decision tree. Advances in neural information processing systems, 30, 2017.

[35] Fabian Pedregosa, Gaël Varoquaux, Alexandre Gramfort, Vincent Michel, Bertrand Thirion, Olivier Grisel, Mathieu Blondel, Peter Prettenhofer, Ron Weiss, Vincent Dubourg, et al. Scikit-learn: Machine learning in python. the Journal of machine Learning research, 12:2825–2830, 2011.

[36] Takuya Akiba, Shotaro Sano, Toshihiko Yanase, Takeru Ohta, and Masanori Koyama. Optuna: A next-generation hyperparameter optimization framework. In Proceedings of the 25th ACM SIGKDD international conference on knowledge discovery & data mining, pages 2623–2631, 2019.

[37] Samuel W Fenske, Alec Peltekian, Mengjia Kang, Nikolay S Markov, Mengou Zhu, Kevin Grudzinski, Melissa J Bak, Anna Pawlowski, Vishu Gupta, Yuwei Mao, et al. Developing and validating machine learning models to predict next-day extubation. Scientific Reports, 15(1): 27552, 2025.

[38] Scott M Lundberg and Su-In Lee. A unified approach to interpreting model predictions. Advances in neural information processing systems, 30, 2017.

[39] Ewa Wielosz, Magdalena Dryglewska, and Maria Majdan. Clinical consequences of the presence of anti-rna pol iii antibodies in systemic sclerosis. Advances in Dermatology and Allergol-ogy/Postffpy Dermatologii i Alergologii, 37(6):909–914, 2020.

[40] Bernadette Lynch, Henry Penn, Jennifer Harvey, Aine Burns, and Christopher P Denton. The prognosis of scleroderma renal crisis in rna-polymerase iii antibody (ara) positive compared to ara negative patients. In ARTHRITIS AND RHEUMATISM, volume 65, pages S1106–S1107. WILEY-BLACKWELL, 2013.

[41] C Bruni, L Petelytska, A Velauthapillai, L Tofani, P Carreira, G Cuomo, E Hachulla, I Castellví, R Bečvář, A Balbir-Gurman, et al. Pos0235 new onset of interstitial lung disease in systemic sclerosis: Clinical course and outcomes from a eustar database analysis. Annals of the Rheumatic Diseases, 83:337–338, 2024.

[42] Giuseppe Murdaca, Simone Caprioli, Alessandro Tonacci, Lucia Billeci, Monica Greco, Simone Negrini, Giuseppe Cittadini, Patrizia Zentilin, Elvira Ventura Spagnolo, and Sebastiano Gangemi. A machine learning application to predict early lung involvement in scleroderma: A feasibility evaluation. Diagnostics, 11(10):1880, 2021.

[43] Guillaume Chassagnon, Maria Vakalopoulou, Alexis Régent, Evangelia I Zacharaki, Galit Aviram, Charlotte Martin, Rafael Marini, Norbert Bus, Naïm Jerjir, Arsène Mekinian, et al. Deep learning–based approach for automated assessment of interstitial lung disease in systemic sclerosis on ct images. Radiology: Artificial Intelligence, 2(4):e190006, 2020.

[44] Janine Schniering, Malgorzata Maciukiewicz, Hubert S Gabrys, Matthias Brunner, Christian Blüthgen, Chantal Meier, Sophie Braga-Lagache, Anne-Christine Uldry, Manfred Heller, Matthias Guckenberger, et al. Computed tomography-based radiomics decodes prognostic and molecular differences in interstitial lung disease related to systemic sclerosis. European Respiratory Journal, 59(5), 2022.

[45] Satoshi Ebata, Ayumi Yoshizaki, Takemichi Fukasawa, Asako Yoshizaki-Ogawa, Yoshihide Asano, Kosuke Kashiwabara, Koji Oba, and Shinichi Sato. Increased red blood cell distribution width in the first year after diagnosis predicts worsening of systemic sclerosis-associated interstitial lung disease at 5 years: a pilot study. Diagnostics, 11(12):2274, 2021.

[46] Shenyun Shi, Ling Chen, Xianhua Gui, Lulu Chen, Xiaohua Qiu, Min Yu, and Yonglong Xiao. Association of red blood cell distribution width levels with connective tissue disease-associated interstitial lung disease (ctd-ild). Disease Markers, 2021(1):5536360, 2021.

[47] Brody H Foy, Rachel Petherbridge, Maxwell T Roth, Cindy Zhang, Daniel C De Souza, Christopher Mow, Hasmukh R Patel, Chhaya H Patel, Samantha N Ho, Evie Lam, et al. Haematological setpoints are a stable and patient-specific deep phenotype. Nature, 637(8045):430–438, 2025.

[48] Lauren Rose, Kurt W Prins, Stephen L Archer, Marc Pritzker, E Kenneth Weir, Jeffrey R Misialek, and Thenappan Thenappan. Survival in pulmonary hypertension due to chronic lung disease: influence of low diffusion capacity of the lungs for carbon monoxide. The Journal of heart and lung transplantation, 38(2):145–155, 2019.

[49] Alec K Peltekian, Karolina Senkow, Gorkem Durak, Kevin M Grudzinski, Bradford C Bemiss, Jane E Dematte, Carrie Richardson, Nikolay S Markov, Mary Carns, Kathleen Aren, et al. Imaging-based mortality prediction in patients with systemic sclerosis. In International Workshop on PRedictive Intelligence In MEdicine, pages 14–23. Springer, 2025.

[50] Alec K Peltekian, Halil Ertugrul Aktas, Gorkem Durak, Kevin Grudzinski, Bradford C Bemiss, Carrie Richardson, Jane E Dematte, GR Budinger, Anthony J Esposito, Alexander Misharin, et al. Ren: Anatomically-informed mixture-of-experts for interstitial lung disease diagnosis. *arXiv preprint arXiv:2510.04923*, 2025.

[51] Brendan G Cooper, Janet Stocks, Graham L Hall, Bruce Culver, Irene Steenbruggen, Kim W Carter, Bruce Robert Thompson, Brian L Graham, Martin R Miller, Gregg Ruppel, et al. The global lung function initiative (gli) network: bringing the world’s respiratory reference values together. Breathe, 13(3):e56–e64, 2017.

[52] Cole Bowerman, Nirav R Bhakta, Danny Brazzale, Brendan R Cooper, Julie Cooper, Laura Gochicoa-Rangel, Jeffrey Haynes, David A Kaminsky, Le Thi Tuyet Lan, Refiloe Masekela, et al. A race-neutral approach to the interpretation of lung function measurements. American journal of respiratory and critical care medicine, 207(6):768–774, 2023.

[53] National Kidney Foundation. Estimated glomerular filtration rate (egfr). https://www.kidney.org/kidney-topics/estimated-glomerular-filtration-rate-egfr, 2025. Accessed: 2025-10-09.

