## Supplemental Methods/Figures for "Machine Learning Analysis of Electronic Health Records Identifies Interstitial Lung Disease and Predicts Mortality in Patients with Systemic Sclerosis"

### Supplementary Methods and Materials

#### Supplementary Contents

#### Feature Extraction

##### Categorical Variables

- **Demographics:** Ethnicity, sex, and race were extracted from Northwestern University's clinical trial management system: StudyTracker. Ethnicity and sex were encoded as binary variables, while race was represented using multi-category groupings, including multiracial classifications.

- **Subtype:** Systemic sclerosis (SSc) subtypes—limted cutaneous (lcSSc), diffuse cutaneous (dcSSc), and sine scleroderma (SSS)—were derived from Registry records. Diagnoses outside these categories (e.g., morphea, very early diagnosis of SSc, mixed or undifferentiated connective tissue disease, overlap syndromes) were manually reviewed and adjudicated into the three SSc subtypes when appropriate, based on established classification criteria [27, 28].

- **Smoking Status:** This feature was extracted from REDCap and categorized as "current", "former", or "never" smoker. Exposure history was measured in pack-years as a continuous variable, calculated as:

$$\text{Pack years} = \frac{\text{cigarettes per day}}{20} \times \text{years smoked.}$$

- **Autoantibodies:** Anti-topoisomerase I (Scl-70), anticentromere (ACA), and anti-RNA polymerase III (RNA Pol III) titer positivities were extracted from laboratory reports and antinuclear antibody (ANA) pattern analyses (in the case of ACA). If a patient tested positive for any autoantibody, missing values for the remaining antibodies were imputed as negative. Remaining missing values were adjudicated by clinician review.

- **Pulmonary Hypertension (PH):** PH status was derived from right heart catheterization (RHC) measurements, including mean pulmonary arterial pressure (mPAP), pulmonary capillary wedge pressure (PCWP), and pulmonary vascular resistance (PVR), calculated as:

$$\text{PVR} = \frac{\text{mPAP - PCWP Mean}}{\text{Cardiac Output}}.$$

  Participants were classified into PH subgroups based on mPAP and PVR values:

1. Pre-capillary PH: mPAP $>$ 20 mmHg and PVR $>$ 2 Woods unit (WU),

2. Post-capillary PH: mPAP $>$ 20 mmHg and PVR $\leq$ 2 WU, or

3. No PH: mPAP $\leq$ 20 mmHg.

For longitudinal classification:

– If a patient had initial "No PH" measurements but later developed PH, the patient was labeled as having PH at the time of the RHC and only the earliest No PH measurement was retained prior to the PH diagnosis.

– If a patient was diagnosed with pre- or post-capillary PH and subsequently had a RHC without PH, the patient retained the PH label.

– If a patient was diagnosed with PH and had no other subsequent RHC data, the patient retained their PH label longitudinally.

- **Treatment with mycophenolate mofetil (Rx on MMF):** A time-updated variable indicating whether a patient had ever received MMF. The variable was set to 0 until the patient's first documented MMF dispensed, after which it remained 1 for all subsequent timepoints (naïve carry-forward encoding). Only participants with at least two documented MMF prescriptions were labeled as "Rx on MMF" in the analyses.

**Continuous Variables**

- **Demographics:** Age was calculated from date of birth and to event dates.

- **Complete Blood Count Measurements:** This laboratory panel included the following features: mean corpuscular hemoglobin concentration (MCHC), mean corpuscular hemoglobin (MCH), red cell distribution width (RDW), mean corpuscular volume (MCV), hematocrit, platelet count, hemoglobin, red and white blood cell counts, lymphocytes (percent and absolute), basophils (percent and absolute), eosinophils (percent and absolute), mean platelet volume (MPV; percent and absolute), neutrophils (percent and absolute), and monocytes (percent and absolute). Absolute counts (if not reported) were derived using the following formula, and outliers (e.g., implausible values) were removed through expert adjudication:

$$\text{Absolute Count} = \text{Feature} \times \text{White Cell Count} \times 10$$

- **C-Reactive Protein (CRP):** Values were extracted from the electronic health record (EHR).
- **Pulmonary Function Tests (PFTs):** Forced vital capacity (FVC), forced expiratory volume in one second (FEV1), and diffusing capacity of the lung for carbon monoxide (DLCO) were extracted from the Registry, electronic data warehouse (EDW), and external reports from other centers imported into the EHR. PFTs taken less than one month apart were consolidated to retain the most reliable value through expert adjudication. Chart reviews were performed to avoid redundant measurements. Outliers were removed. Values were normalized to values derived from race-neutral equations provided by the Global Lung Function Initiative (GLI) as follows:
  - GLI adjustments: Predicted FVC and FEV1 values were adjusted based on GLI equations [51, 52]. Absolute values were then divided by the GLI-corrected predicted value to obtain percent predicted.
  - Hemoglobin (Hgb) correction for DLCO:

$$\text{Age-Sex Factor} = \begin{cases} 9.38, & \text{if female or age} < 15 \\ 10.22, & \text{otherwise} \end{cases}$$

$$\text{DLCO\_REF\_Hg\_Corrected} = \text{DLCO\_REF} \times \frac{1.7 \times \text{Hemoglobin}}{\text{Age-Sex Factor} + \text{Hemoglobin}}$$

$$\text{DLCO} = \frac{\text{DLCO actual}}{\text{DLCO\_REF\_Hg\_Corrected}} \times 100$$

- **6-Minute Walk Test:** Distance walked in meters was extracted from EHR reports obtained from the EDW.
- **Chemistries:** Values for serum concentrations of creatinine, alkaline phosphatase, alanine aminotransferase (ALT), anion gap, aspartate aminotransferase (AST), bicarbonate, blood

urea nitrogen (BUN), calcium, chloride, potassium, total protein, sodium, and total bilirubin were extracted from EHR reports obtained from the EDW. Glomerular filtration rate (GFR) was also extracted and categorized.

GFR was preprocessed to follow an ordinal scale reflecting the stages of chronic kidney disease (CKD).[53] Estimated GFR values were mapped to an ordinal scale (1–5), with higher values indicating more preserved renal function. Values reported as $> 60$ mL/min/1.73m$^2$ were grouped into a single category due to EHR reporting limitations. Values 60–90 and $> 90$ were combined into the same category since the EHR system reported them as $> 60$.

- **B-type Natriuretic Peptide (BNP) and Modified Rodnan Skin Score (mRSS):** BNP and mRSS were extracted from the EHR, with additional manual review to filter outlier or implausible mRSS values.
- **Vitals:** This included body mass index (BMI), diastolic blood pressure, systolic blood pressure, body surface area (BSA), height, pulse, and weight.

#### Modeling

##### Model Types

We used a range of machine learning (ML) models to address our prediction tasks. Logistic Regression was implemented to leverage linear modeling between features and the log odds of the outcomes. Random Forest Classifier was used as an ensemble approach. Gradient Boosting Machines (GBM), including XGBoost [33] and LightGBM [34], were chosen for their robustness in handling missing data and their ability to capture complex feature interactions. Neural Networks, specifically the Multi-Layer Perceptron (MLP) Classifier from the scikit-learn library [35], were also utilized, employing architectures such as single-layer networks (e.g., fifty nodes) or stacked layers (e.g., thirty and fifteen nodes).

##### Imputation Strategy

Continuous features were organized into quartiles, with missingness encoded using binary mask indicators. Categorical features encoded missing values as an explicit category and were one-hot encoded. Quantile thresholds were derived from the training set and applied consistently across validation and test splits to prevent data leakage.

##### Optimization and Model Selection

Hyperparameter optimization was conducted using Optuna [36], a framework that efficiently explores the hyperparameter space. Each model-task pair underwent up to twenty-five trials to ensure comprehensive tuning, with performance metrics such as area under the curve (AUC) and loss tracked across iterations. Five-fold cross validation was also performed for model selection. Upon averaging the five scores for each trial, along with the standard deviation, we calculated a score using:

$$\text{Score} = \text{Mean} - 0.5 \times \text{Standard Deviation},$$

penalizing models with high cross-validation variance. Final model selection was based on the highest score across all model types.

##### Feature Importance

Feature importance was assessed using SHapley Additive exPlanations (SHAP) analysis [38] for tree-based models and complementary ablation studies by setting features in the test sets to NaN and observing drops in predictive capability. SHAP values quantified feature contributions at the individual prediction level, while ablation analyses measured performance degradation following systematic feature removal using mask-based encoding. Together, these approaches provided an assessment of predictors associated with ILD and related outcomes.

#### Supplementary Figures/Tables

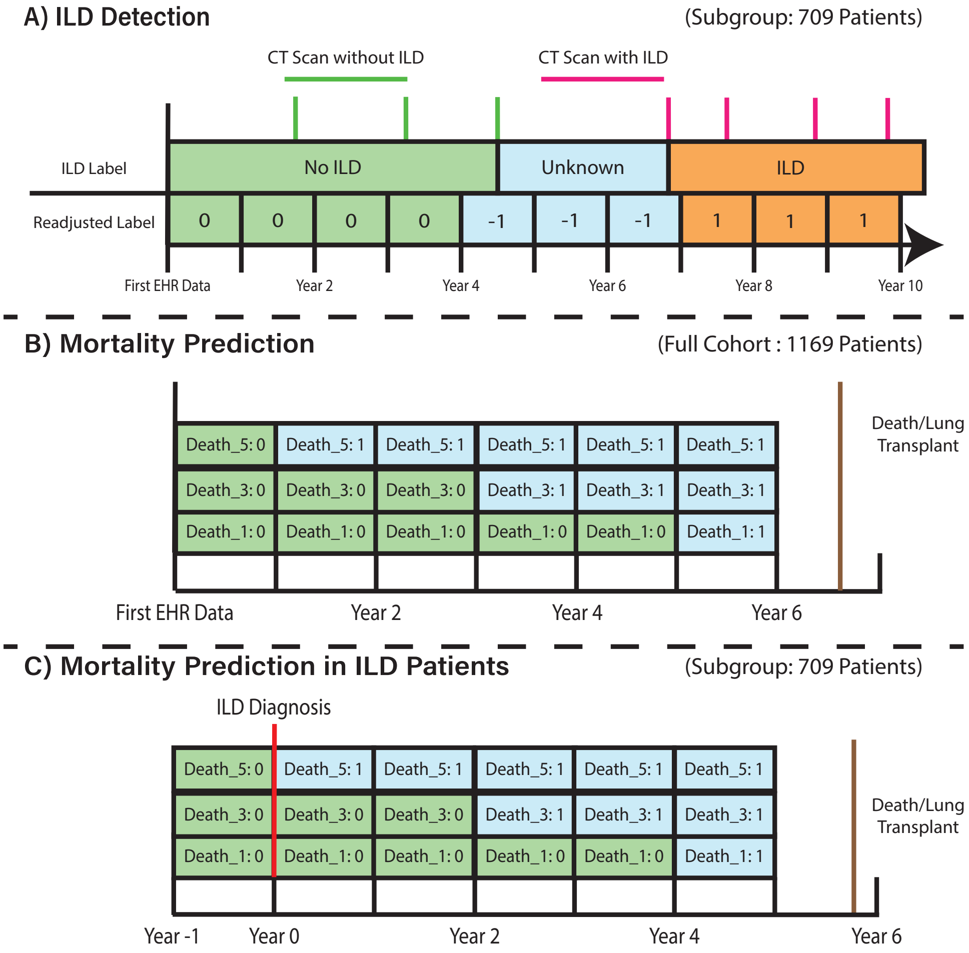

Figure S1: **Labeling strategies and prediction tasks for interstitial lung disease (ILD) detection and mortality modeling.** (**A**) ILD detection timeline based on longitudinal CT imaging. Participants were labeled as 0 (no ILD), -1 (uncertain interval between negative CT and diagnostic CT), or 1 (confirmed ILD). Green markers indicate CT scans without ILD and red markers indicate CT scans confirming ILD. (**B**) Mortality prediction in all participants with systemic sclerosis (SSc), defined using annual prediction bins relative to the time of death or lung transplant. (**C**) Mortality prediction in participants with SSc-associated ILD, with prediction windows beginning one year prior to ILD diagnosis.

Table S1: Cohort Characteristics (CT Subgroups)

| Characteristic | Northwestern Cohort (N = 709) | Yale Cohort (N = 223) |
|---|---|---|
| **Demographics** | | |
| Age at SSc Diagnosis, years, median [Q1, Q3] | 45.1 [36.0, 53.4] | 50.4 [38.2, 61.2] |
| Sex, female, n (%) | 585 (82.5) | 187 (83.9) |
| **Race, n (%)** | | |
| Asian | 26 (3.7) | 3 (1.3) |
| Black | 96 (13.5) | 22 (9.9) |
| White | 567 (80.0) | 164 (73.5) |
| Other | 20 (2.8) | 34 (15.2) |
| **Ethnicity, n (%)** | | |
| Hispanic or Latino | 66 (9.3) | 36 (16.1) |
| Non-Hispanic or Latino | 636 (89.7) | 182 (81.6) |
| Unknown | 7 (1.0) | 5 (2.2) |
| **Tobacco Smoking Status, n (%)** | | |
| Current | 20 (2.8) | 7 (3.1) |
| Former | 163 (23.0) | 30 (13.5) |
| Never | 273 (38.5) | 86 (38.6) |
| Unknown | 253 (35.7) | 100 (44.8) |
| **Alive, n (%)** | 566 (79.8) | 211 (94.6) |
| **Systemic Sclerosis Subtype, n (%)** | | |
| Limited Cutaneous | 377 (53.2) | 157 (70.4) |
| Diffuse Cutaneous | 297 (41.9) | 51 (22.9) |
| Sine Scleroderma | 35 (4.9) | 12 (5.4) |
| **Autoantibodies*, n (%)** | | |
| Scl-70 | 240 (33.9) | 24 (10.8) |
| Centromere | 147 (20.7) | 32 (14.3) |
| RNA polymerase III | 138 (19.5) | 22 (9.9) |
| **ILD Status, n (%)** | | |
| Negative | 255 (36.0) | 105 (47.1) |
| Incident | 34 (4.8) | 17 (7.6) |
| Prevalent | 420 (59.2) | 101 (45.3) |
| Unknown** | | |
| SSc to ILD Onset, years, median [Q1, Q3] | 3.4 [1.2, 7.9] | 2.1 [0.7, 9.2] |
| **CT Chest Imaging** | | |
| CT Chest Images Available, n (%) | 684 (58.5) | 223 (59.3) |
| Number of CTs per Participant, median [Q1, Q3] | 2.0 [1.0, 5.0] | 2.0 [1.0, 4.0] |
| **Pulmonary Function Tests (PFTs)** | | |
| PFT Available, n (%) | 655 (92.4) | 195 (87.8) |
| Number of PFTs per Participant, median [Q1, Q3] | 5.0 [2.0, 10.0] | 5.0 [2.0, 9.0] |
| FVC (% predicted) | 79.3 [64.2, 93.8] | 91.8 [68.7, 105.0] |
| FEV1 (% predicted) | 78.1 [64.6, 90.9] | 86.6 [69.4, 101.2] |
| DLCO (% predicted) | 60.6 [46.8, 74.1] | 64.0 [48.5, 80.0] |
| **Pulmonary Hypertension (PH) Status†, n (%)** | | |
| No PH | 90 (12.7) | 4 (1.8) |
| Pre-Capillary PH | 112 (15.8) | 7 (3.2) |
| Post-Capillary PH | 64 (9.0) | 3 (1.4) |
| Unknown | 443 (62.5) | 208 (93.7) |

*Participants can be negative for all three major SSc-associated autoantibodies or have missing data for specific autoantibodies (See **Supplemental Section 1** for further details).

**Participants were labeled "unknown" if CT reports were available but expert consensus could not be reached.

†Pulmonary Hypertension definitions are based on right heart catheterization data. Participants were categorized as "no PH", "pre-capillary PH", or "post-capillary PH" using mean pulmonary arterial pressure and pulmonary vascular resistance values.

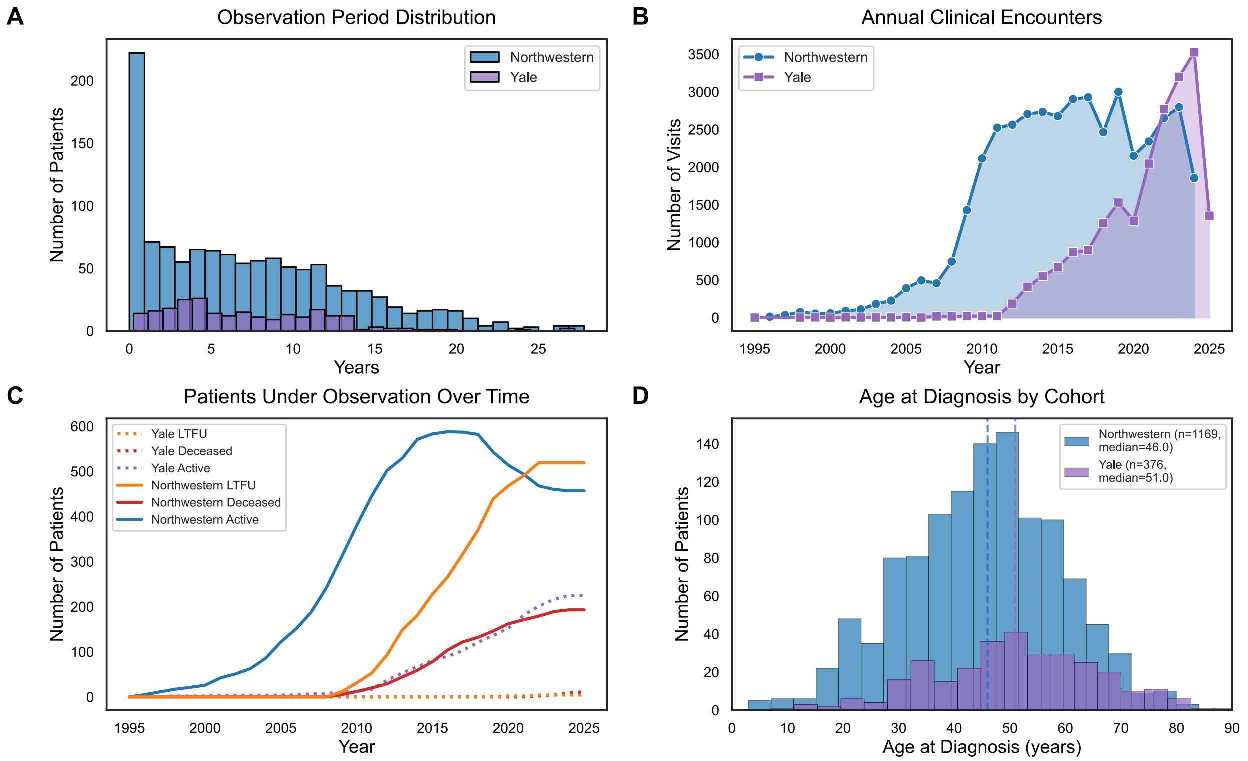

Figure S2: **Longitudinal characteristics of the Northwestern and Yale cohorts.** (**A**) Distribution of participant follow-up duration, binned by years of observation. (**B**) Annual number of recorded clinical encounters over time. (**C**) Cumulative number of participants under observation over time, stratified by active follow-up, loss to follow up, and death. (**D**) Distribution of age at systemic sclerosis diagnosis for participants in each cohort.

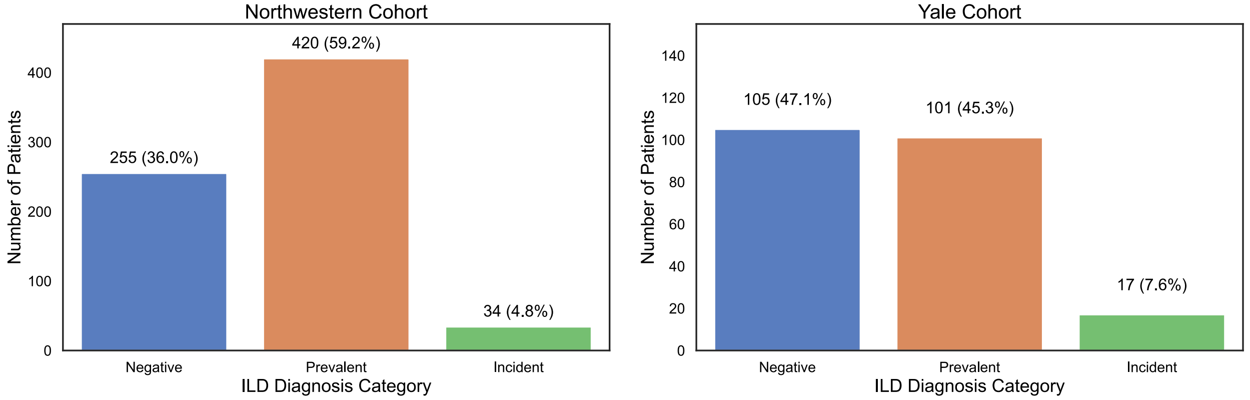

Figure S3: **Distribution of interstitial lung disease (ILD) diagnosis categories in the Northwestern and Yale cohorts.** (**A**) Northwestern cohort illustrating the proportion of participants classified as ILD-negative, ILD-prevalent, or ILD-incident based on longitudinal CT imaging. (**B**) Yale cohort illustrating the corresponding distribution of ILD diagnosis categories.

**(A) Northwestern Cohort.**

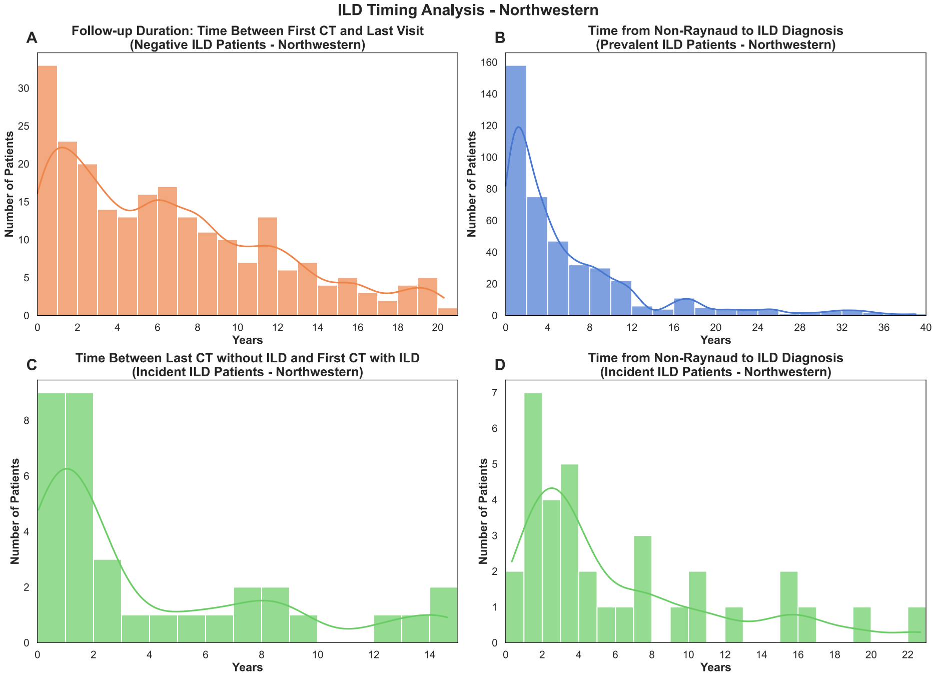

**(B) Yale Cohort.**

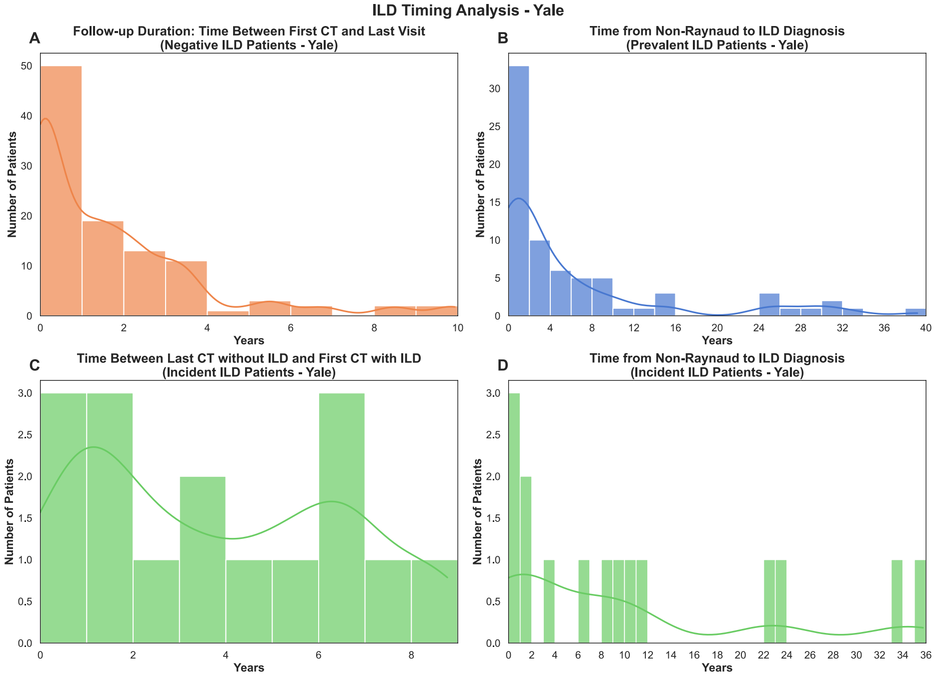

Figure S4: **Timing of ILD onset in the Northwestern and Yale cohorts.** (A) Northwestern cohort and (B) Yale cohort. Panels depict follow-up duration among ILD-negative participants, time from first non-Raynaud symptom to ILD diagnosis in prevalent ILD, time between last normal CT and first ILD-positive CT in incident ILD, and time from first non-Raynaud symptom to ILD diagnosis in incident ILD.

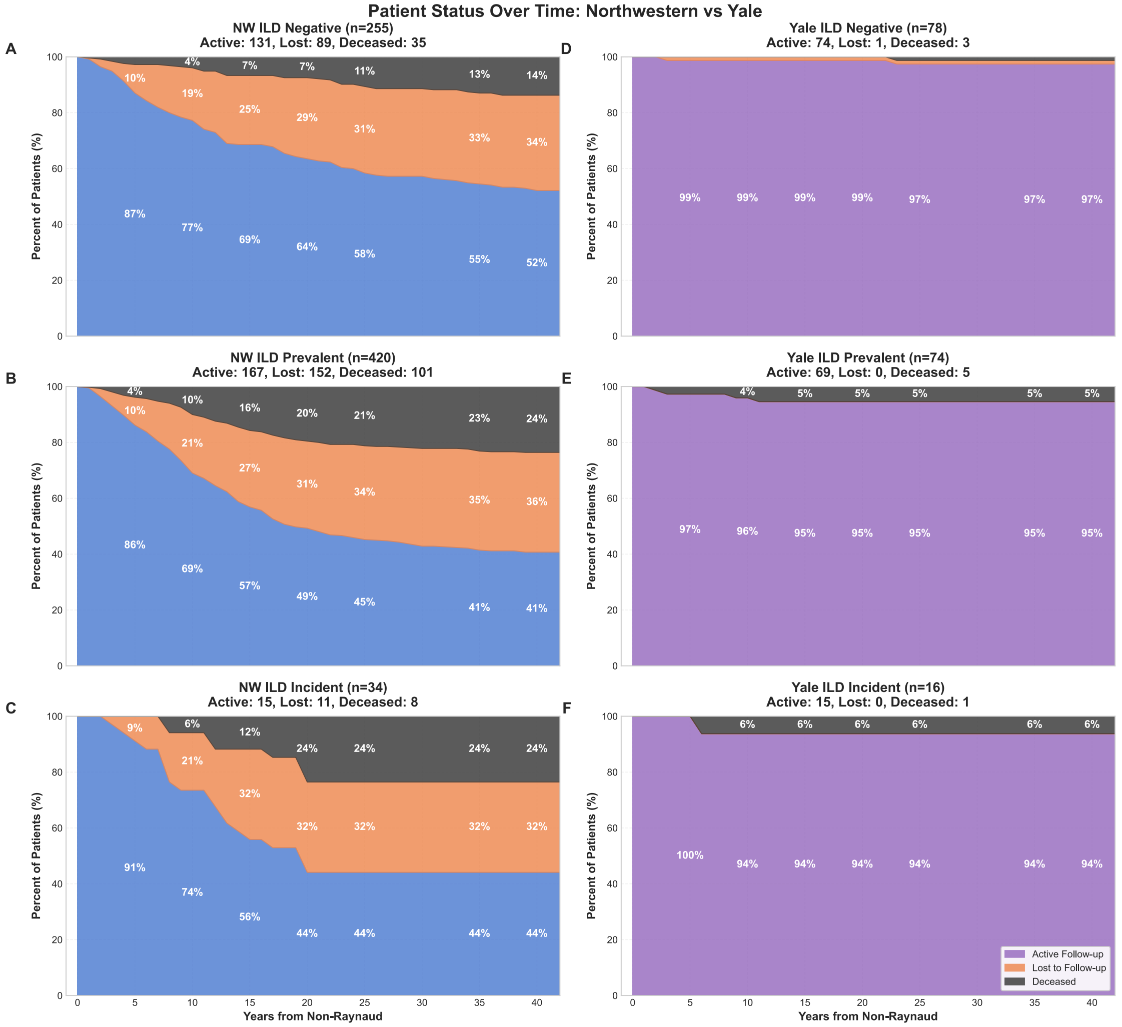

Figure S5: **Survival trajectories stratified by ILD diagnosis category in the Northwestern and Yale cohorts.** (A–C) Northwestern cohort and (D–F) Yale cohort. Panels display the proportion of participants under active follow-up, lost to follow-up, or deceased over time since systemic sclerosis onset, stratified by ILD diagnosis category (negative, prevalent, incident). Loss to follow-up was defined as the absence of recorded clinical measurements for at least two years.

#### (A) Northwestern Cohort.

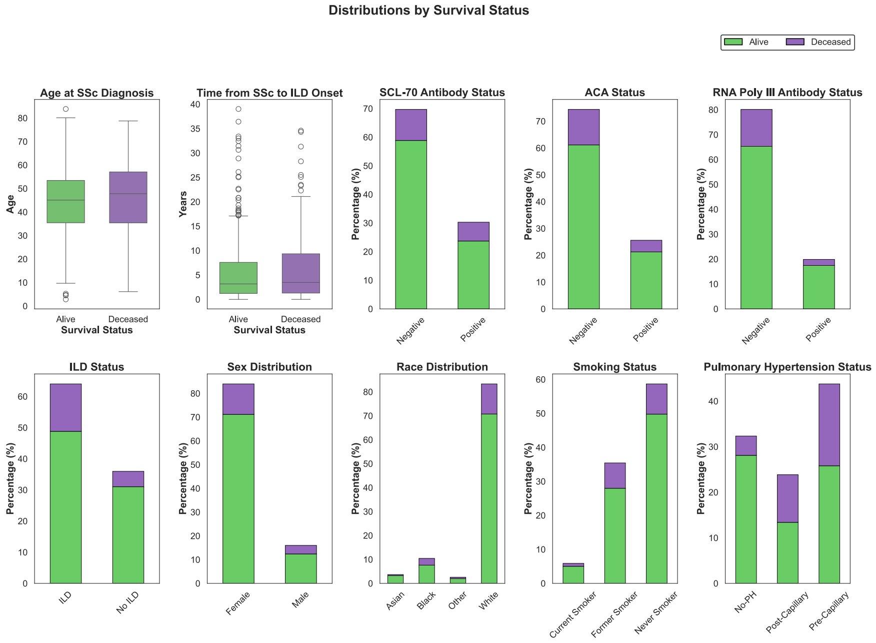

Figure S6: **Mortality distribution across clinical and demographic characteristics in participants with SSc-ILD (Northwestern cohort).** Bar plots display mortality proportions stratified by autoantibody status, ILD status, sex, and pulmonary hypertension classification within the Northwestern cohort.

#### (B) Yale Cohort.

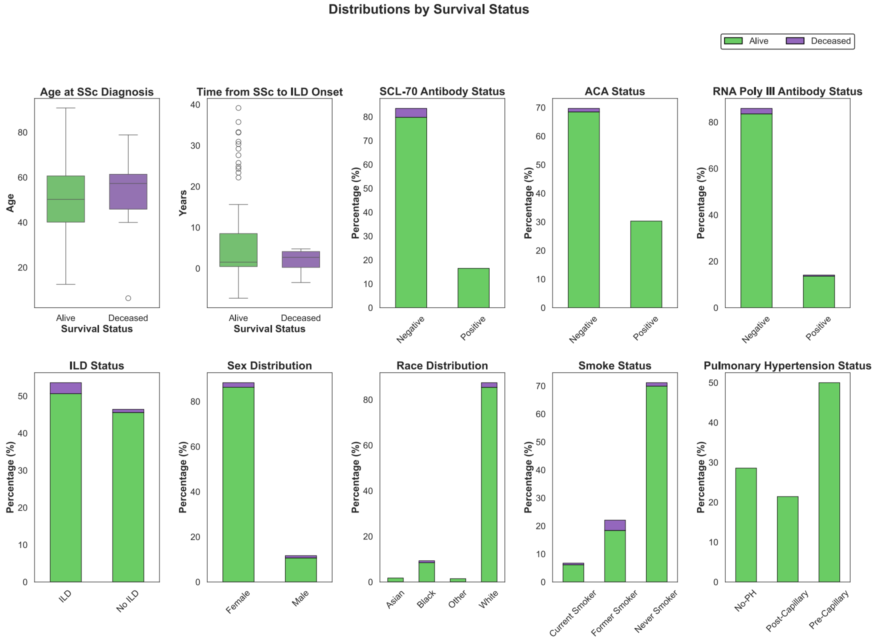

Figure S6: **Mortality distribution across clinical and demographic characteristics in participants with SSc-ILD (continued).** Yale cohort results extending the analysis shown in Figure S6A.

#### (A) Northwestern Cohort.

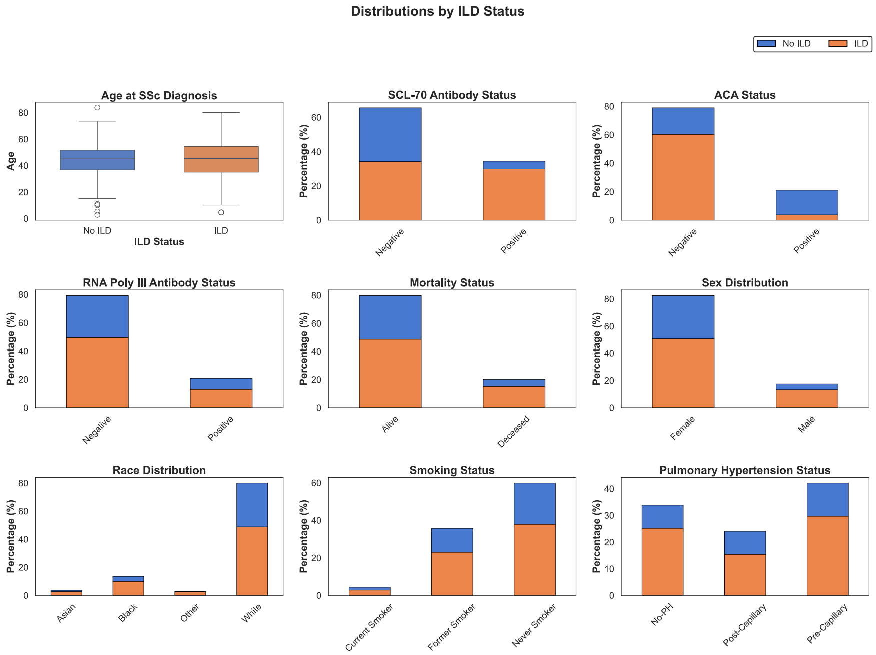

Figure S7: **Distribution of ILD diagnosis across patient characteristics in the Northwestern cohort.** Panels show ILD prevalence stratified by demographic variables, autoantibody status, pulmonary hypertension classification, and age at systemic sclerosis diagnosis.

#### (B) Yale Cohort.

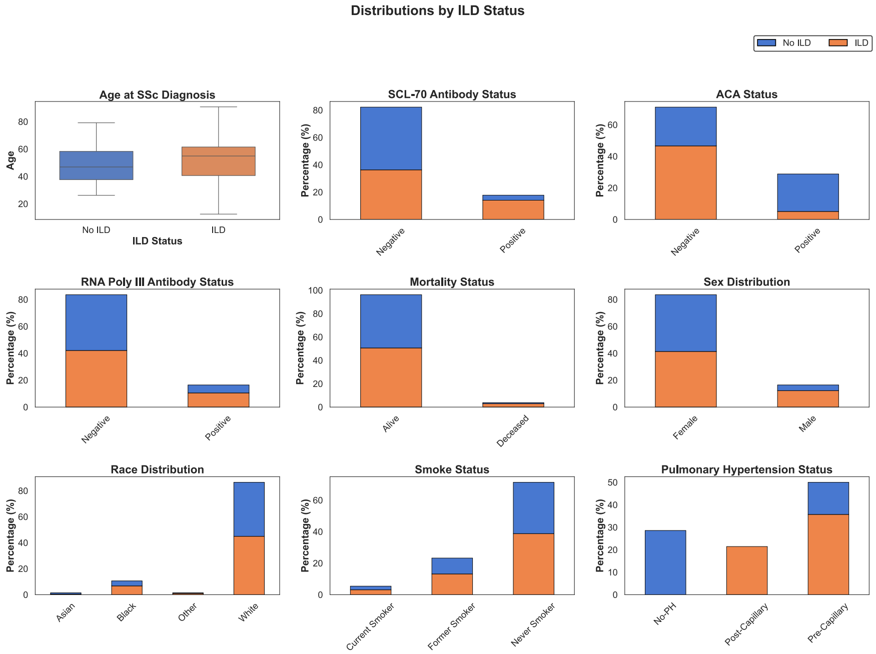

Figure S7: **Distribution of ILD diagnosis across patient characteristics (continued).** Yale cohort results extending the analysis shown in Figure S7A.

##### (A) Northwestern Cohort.

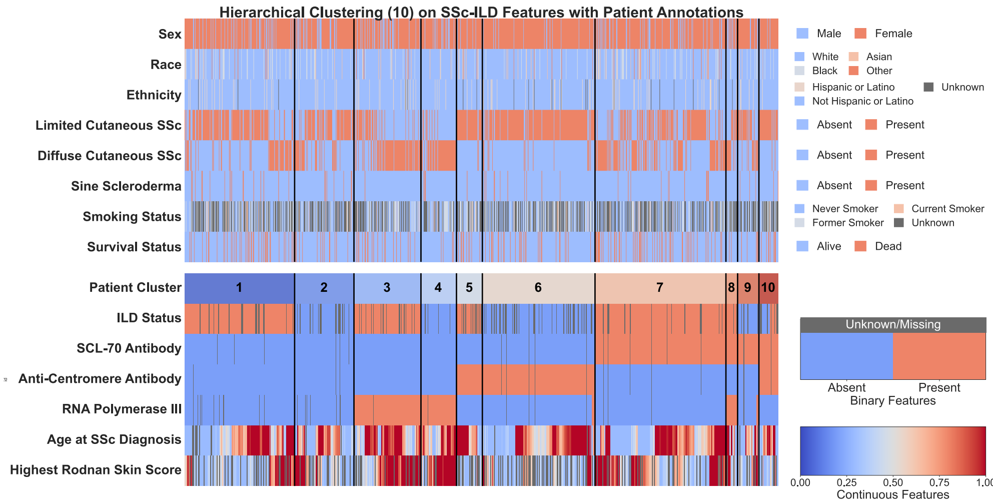

##### (B) Yale Cohort.

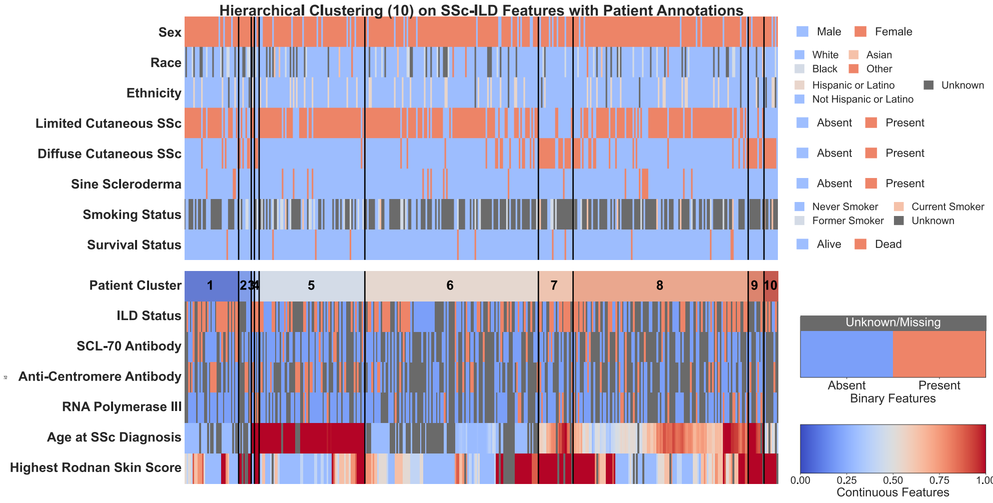

Figure S8: **Hierarchical clustering of systemic sclerosis participants in the Northwestern and Yale cohorts.** Each column represents an individual participant. Top annotations display demographic and clinical characteristics not used for clustering. Bottom panels show hierarchical clustering results (k = 10) derived from longitudinal clinical features.

#### (A) Northwestern Cohort.

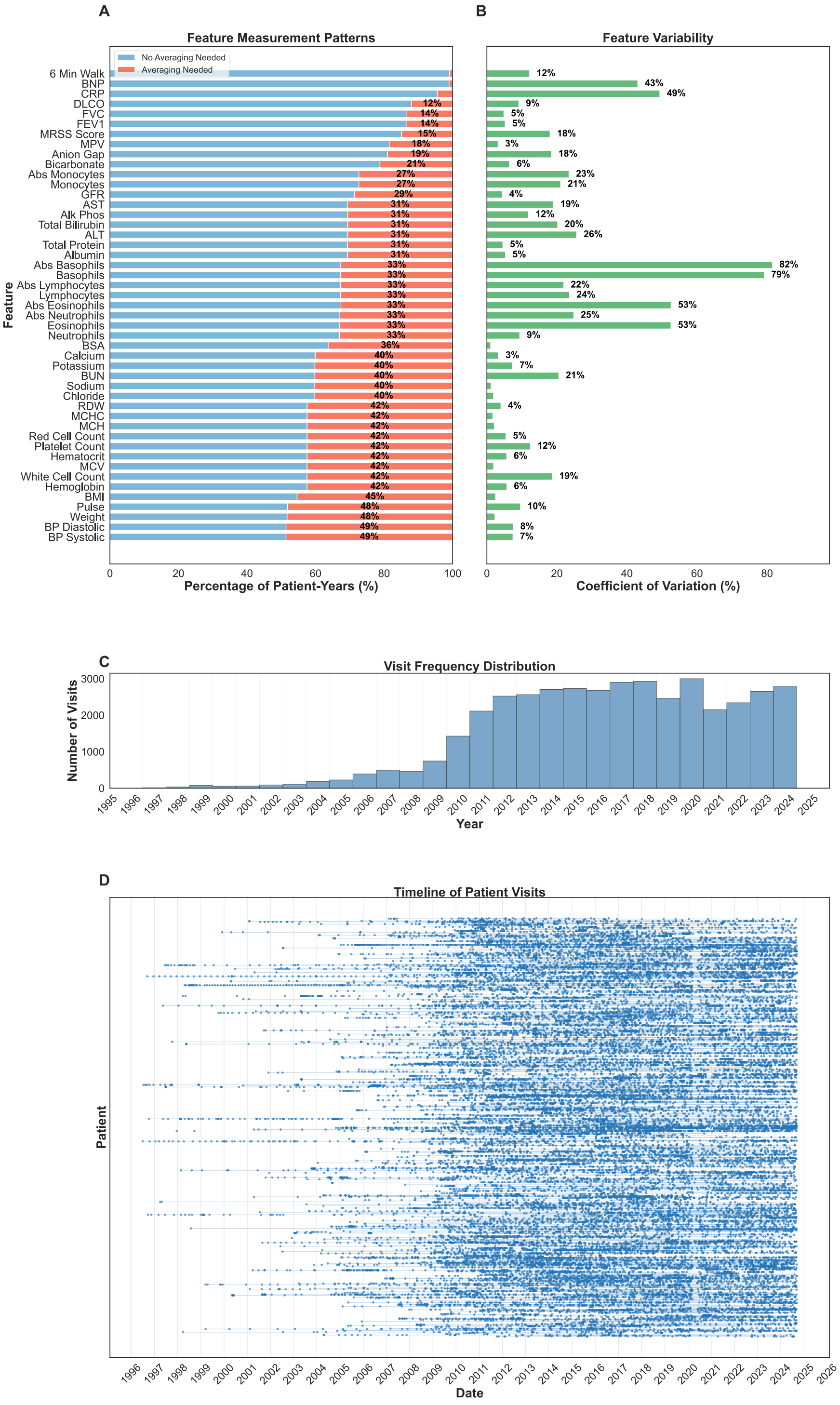

Figure S9: **Patient visit patterns and feature measurement variability in the Northwestern cohort.** (A) Frequency of feature measurements per patient-year. (B) Mean coefficient of variation for features measured multiple times. (C) Distribution of visit frequency across the study period. (D) Longitudinal visit timelines for all participants.

**(B) Yale Cohort.**

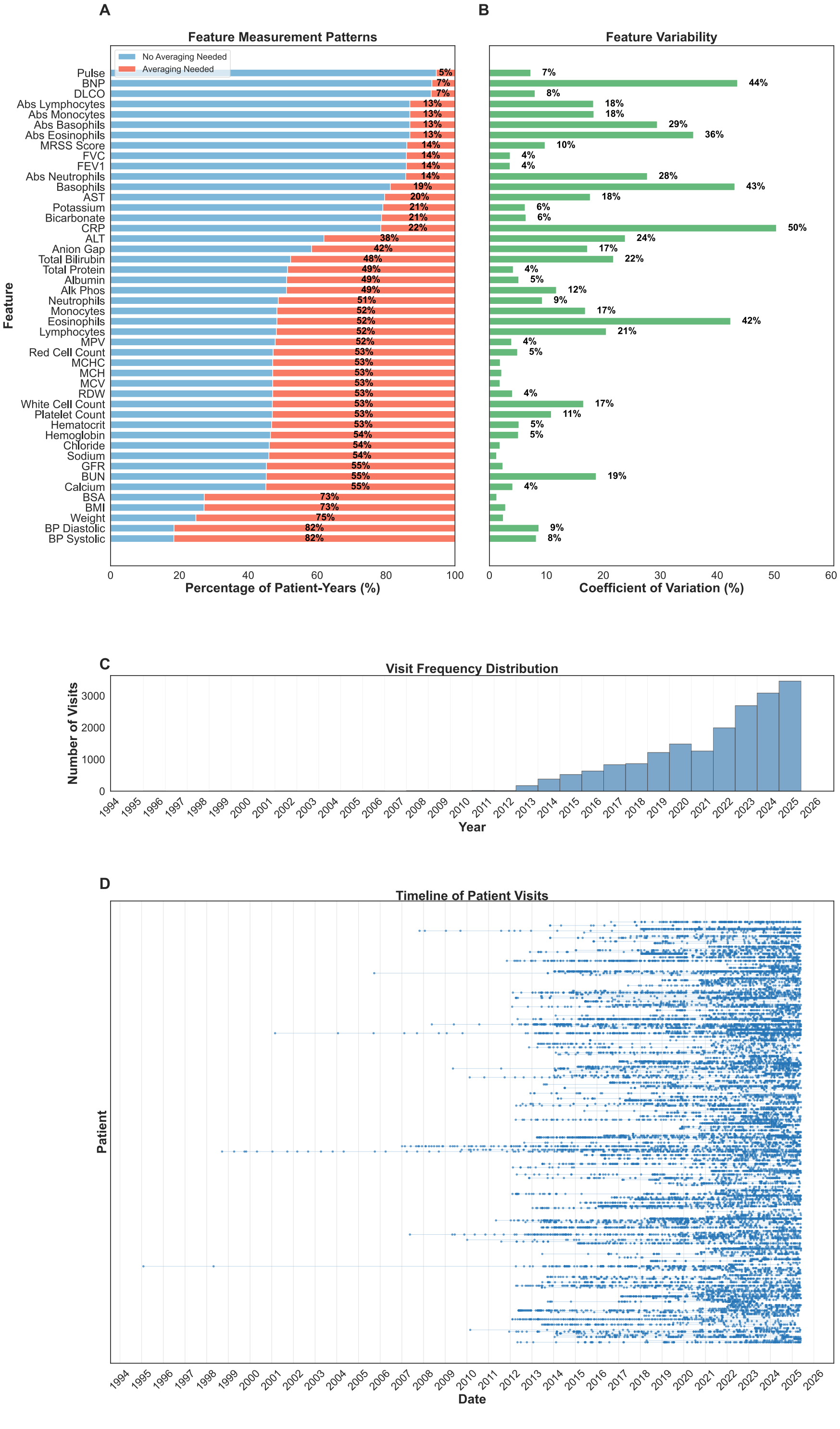

Figure S9: **Patient visit patterns and feature measurement variability (continued).** Yale cohort results extending the analysis shown in Figure S9A.

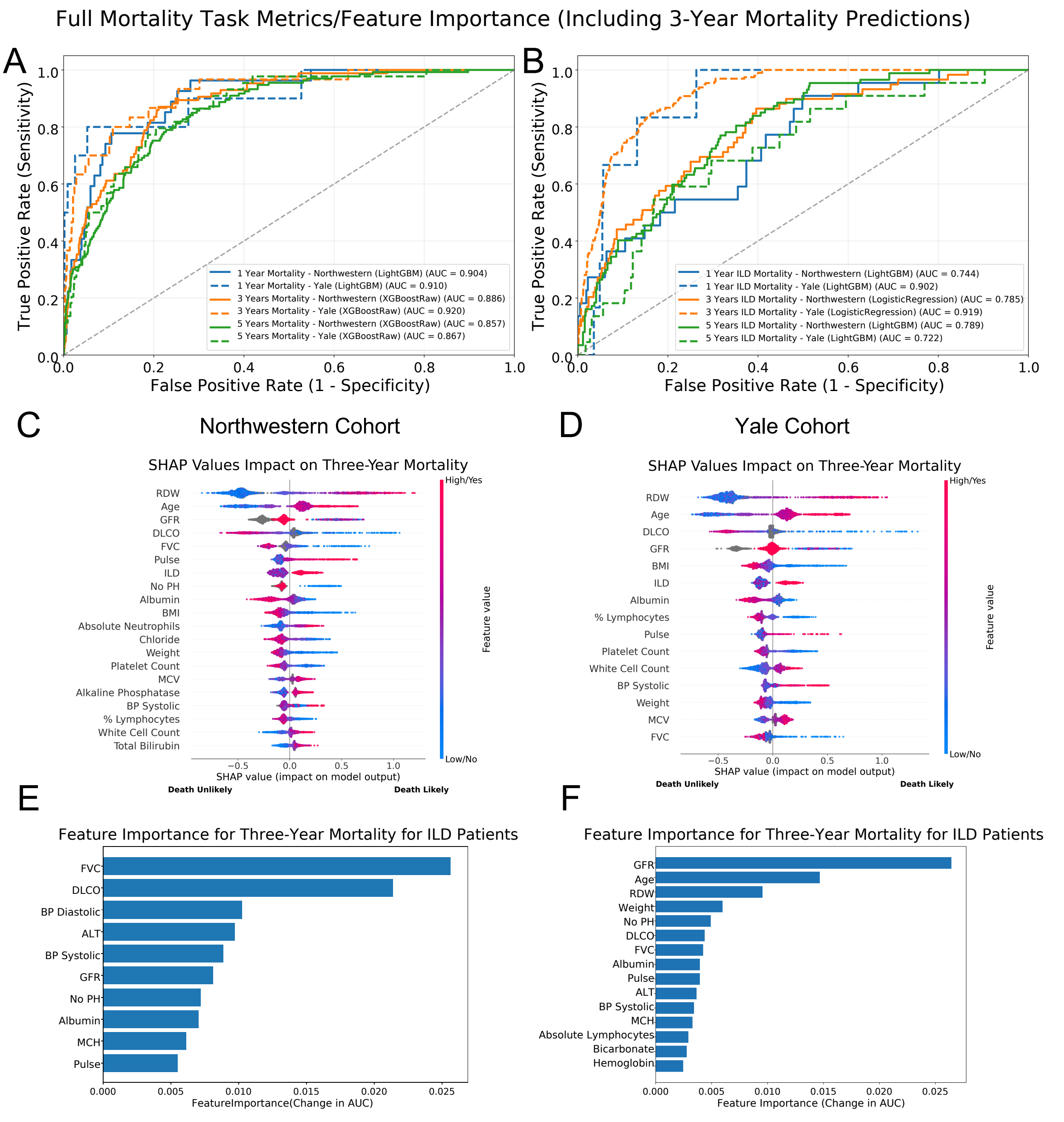

Figure S10: **Model performance and feature importance for three-year mortality prediction in participants with SSc-ILD.** (A,B) ROC curves for mortality prediction in the Northwestern and Yale cohorts. (C,D) SHAP feature importance for LightGBM models. (E,F) Feature importance for logistic regression models trained on the same data.

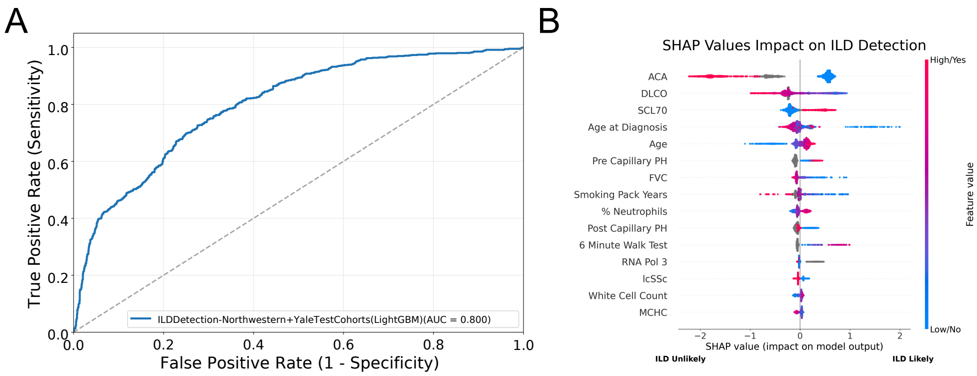

Figure S11: **Model performance and feature importance for ILD detection using combined cohorts.** (A) ROC curve showing performance of the LightGBM model for ILD detection. (B) SHAP summary plot illustrating the relative contribution of top predictors.

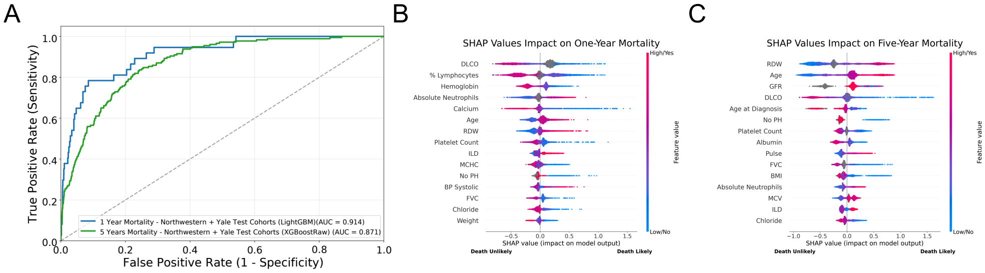

Figure S12: **Model performance and feature importance using combined cohorts for one-year and five-year mortality prediction in all participants with systemic sclerosis.** (A) ROC curves for one-year and five-year mortality prediction. (B,C) SHAP feature importance plots highlighting predictors contributing to mortality risk.

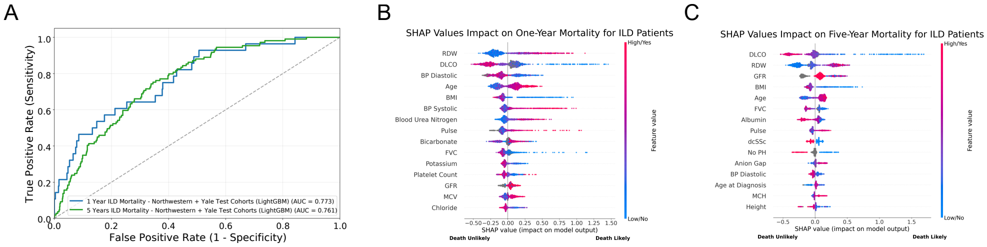

Figure S13: **Model performance and feature importance using combined cohorts for one-year and five-year mortality prediction in participants with SSc-ILD.** (A) ROC curves for one-year and five-year mortality prediction. (B,C) SHAP feature importance plots showing predictors associated with mortality risk in SSc-ILD.

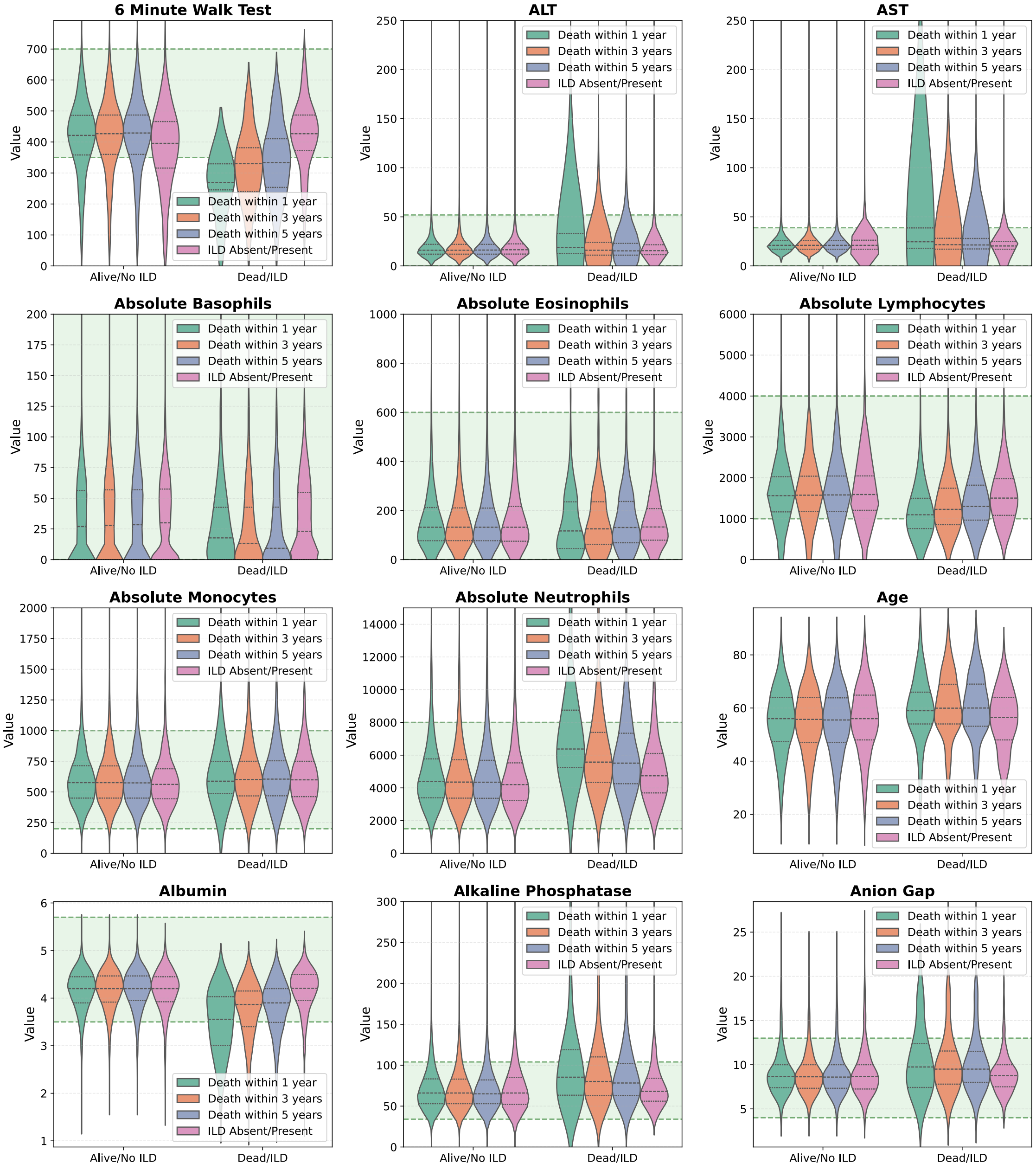

Figure S14: **(A) Distribution of clinical parameters across outcome groups in the Northwestern cohort.** Violin plots display feature distributions stratified by outcome status, with quartile markers and reference ranges where applicable.

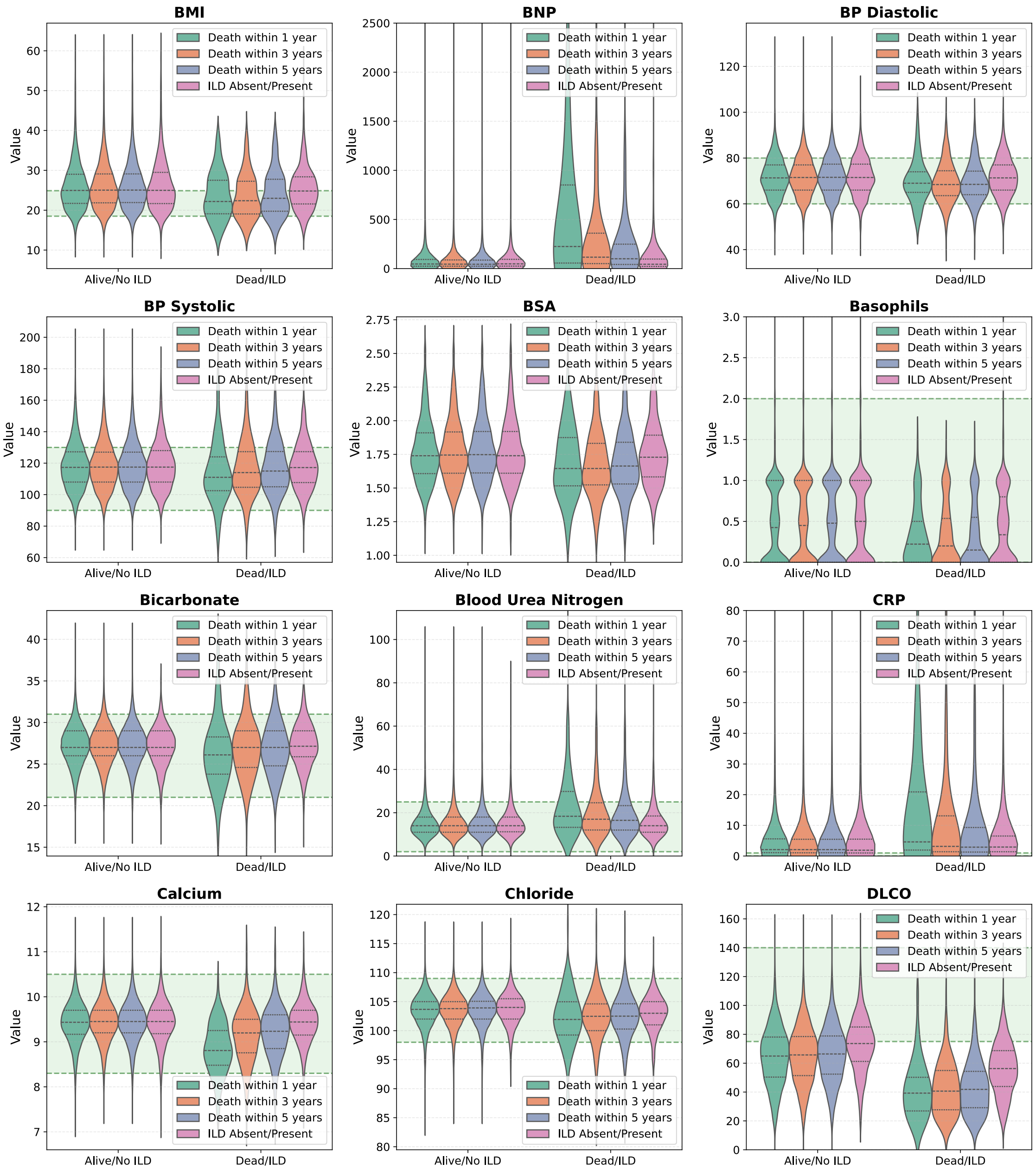

Figure S14: **(Panel B)** Continued from Figure S14A.

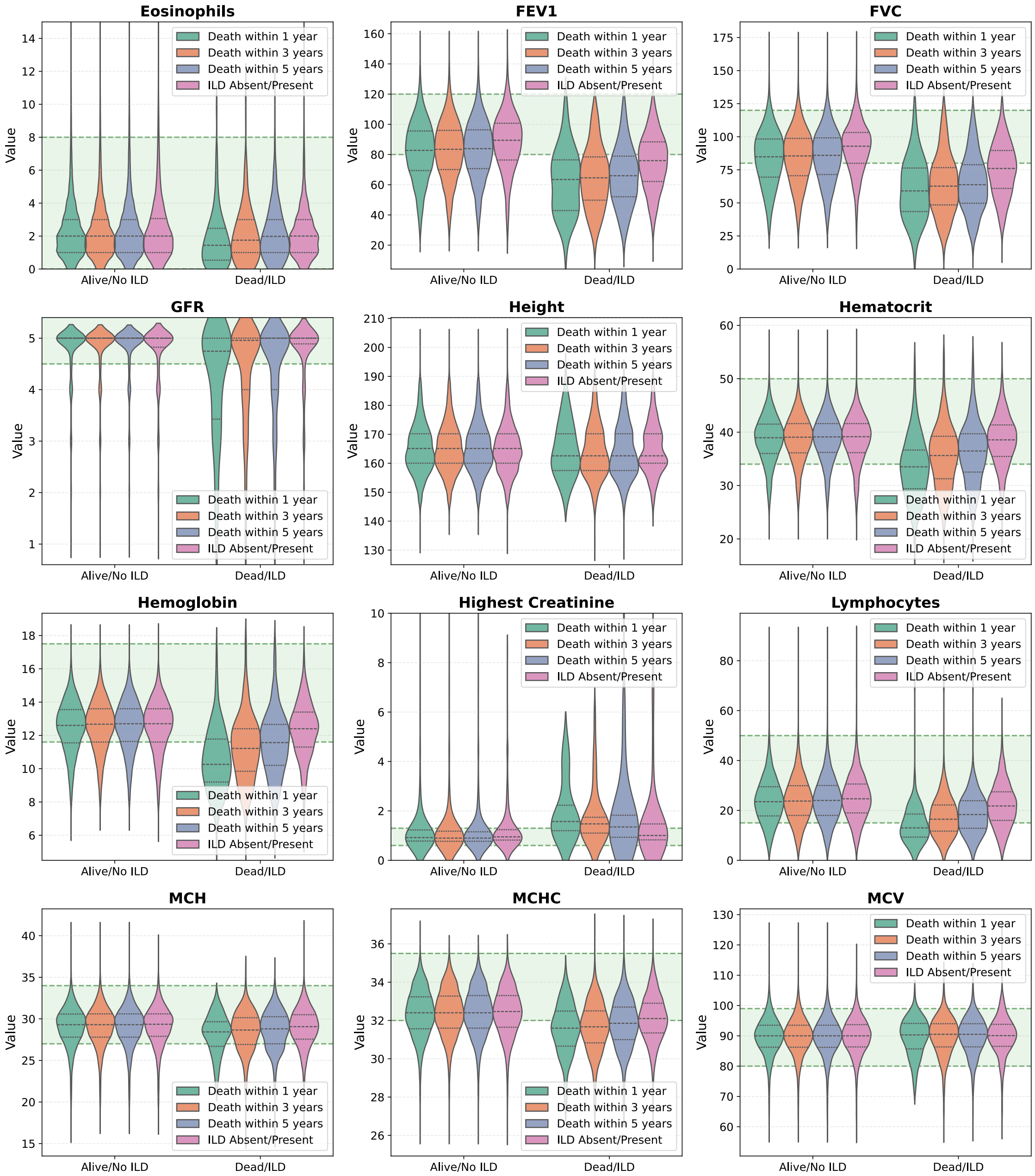

Figure S14: **(Panel C)** Continued from Figure S14B.

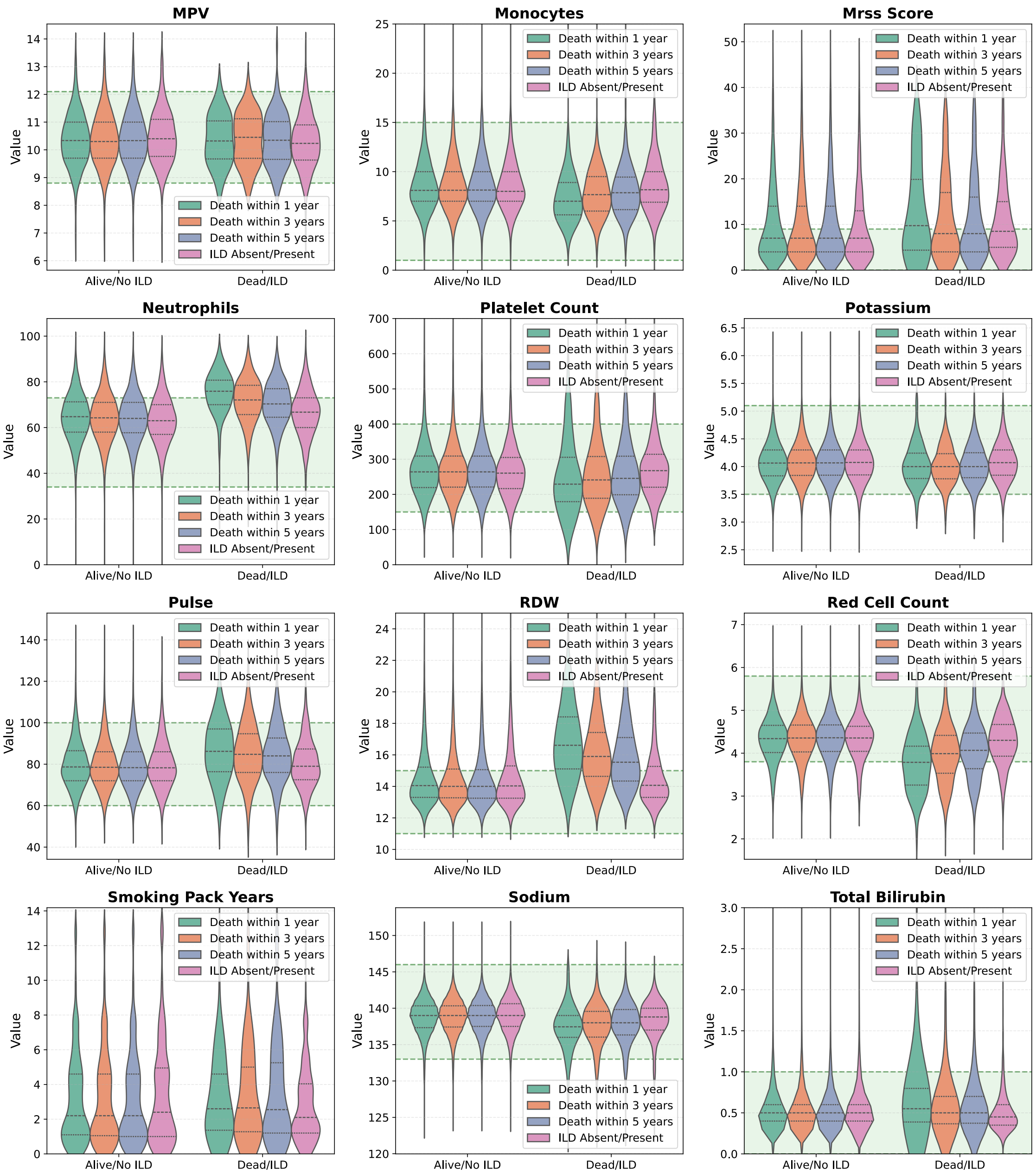

Figure S14: **(Panel D)** Continued from Figure S14C.

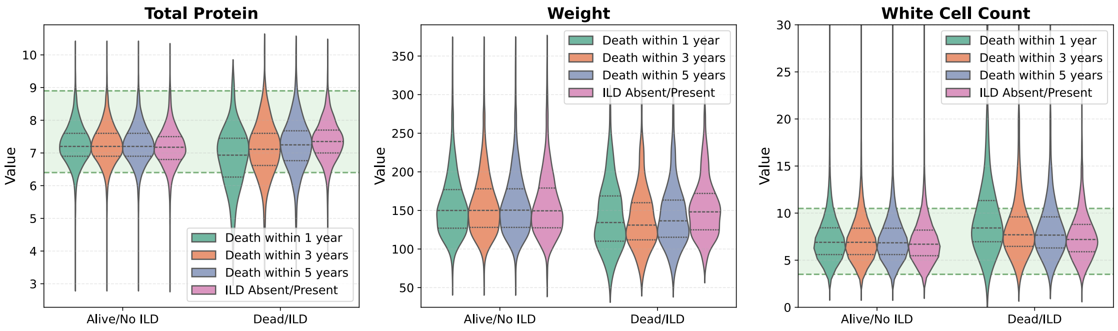

Figure S14: **(Panel E)** Continued from Figure S14D.

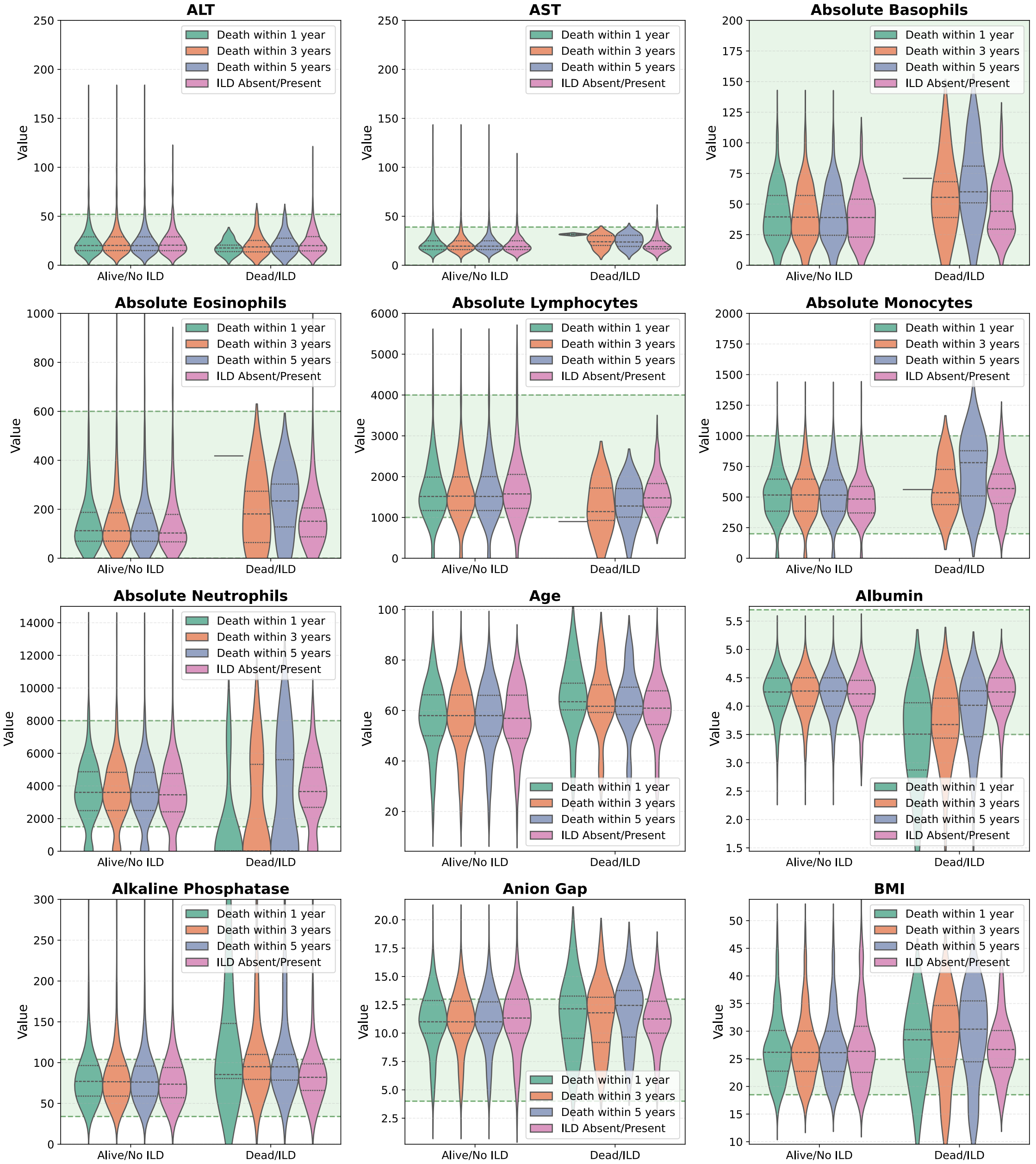

Figure S15: **(A) Distribution of clinical parameters across outcome groups in the Yale cohort.** Violin plots display feature distributions stratified by outcome status, with quartile markers.

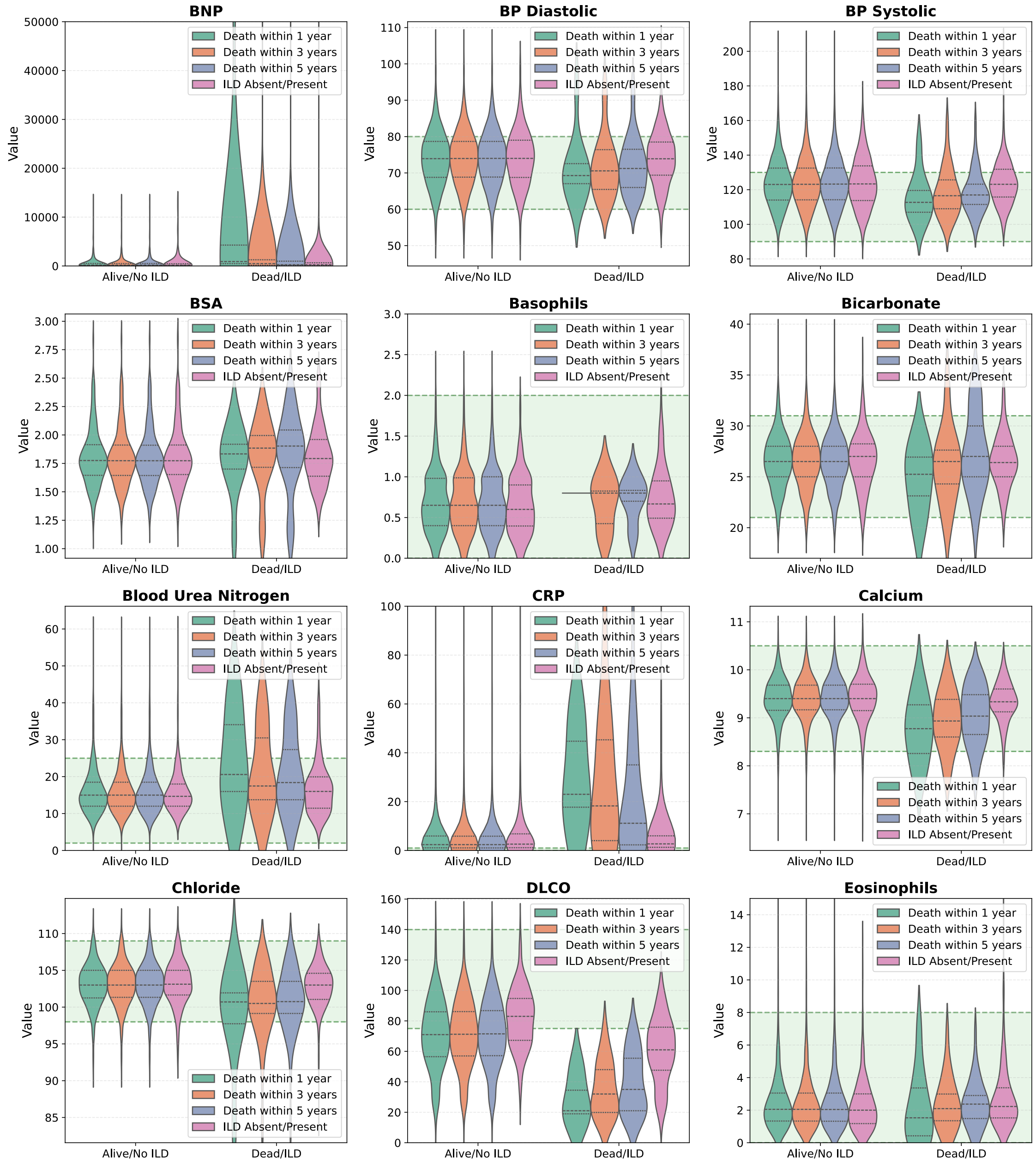

Figure S15: **(Panel B)** Continued from Figure S15A.

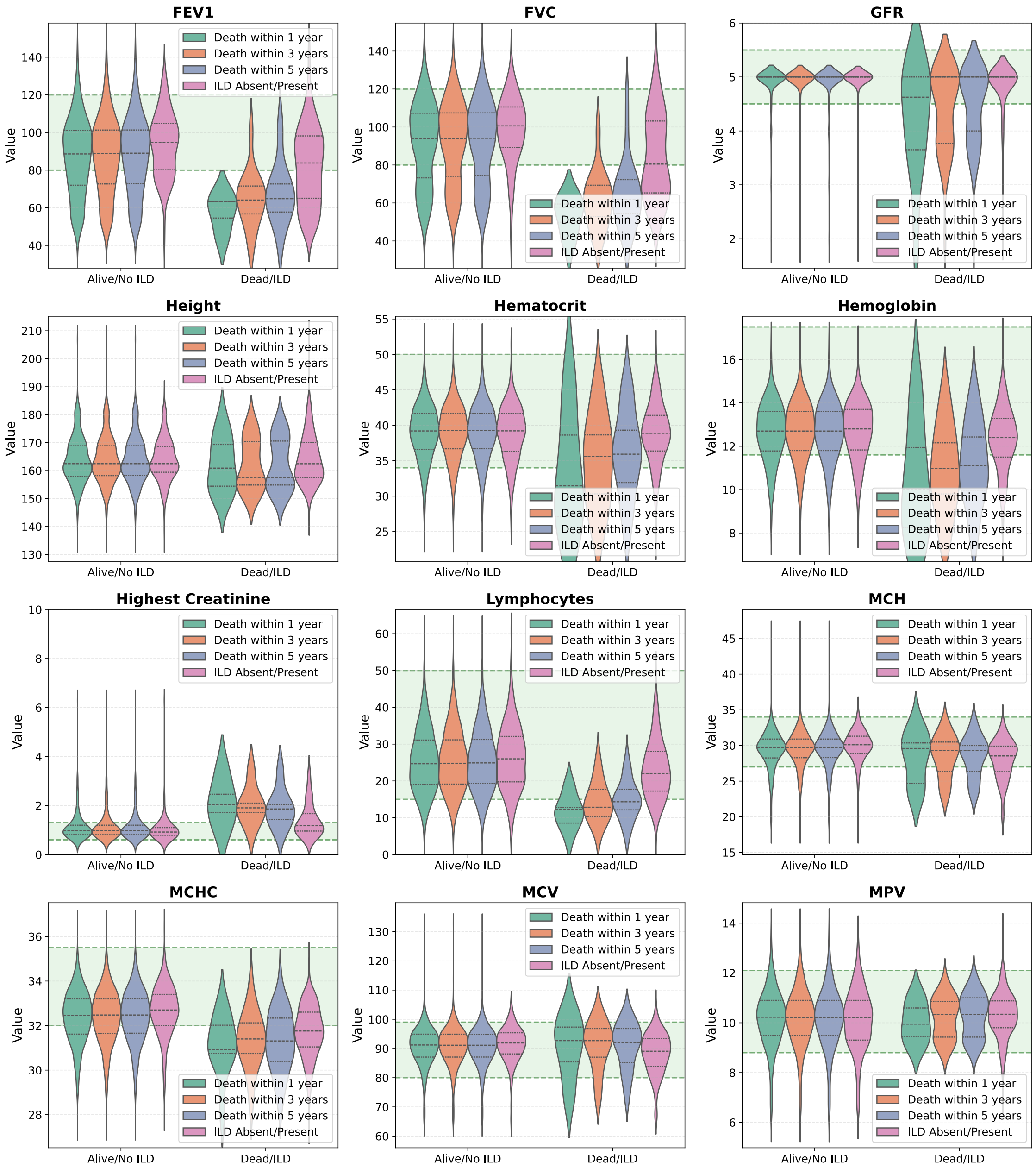

Figure S15: **(Panel C)** Continued from Figure S15B.

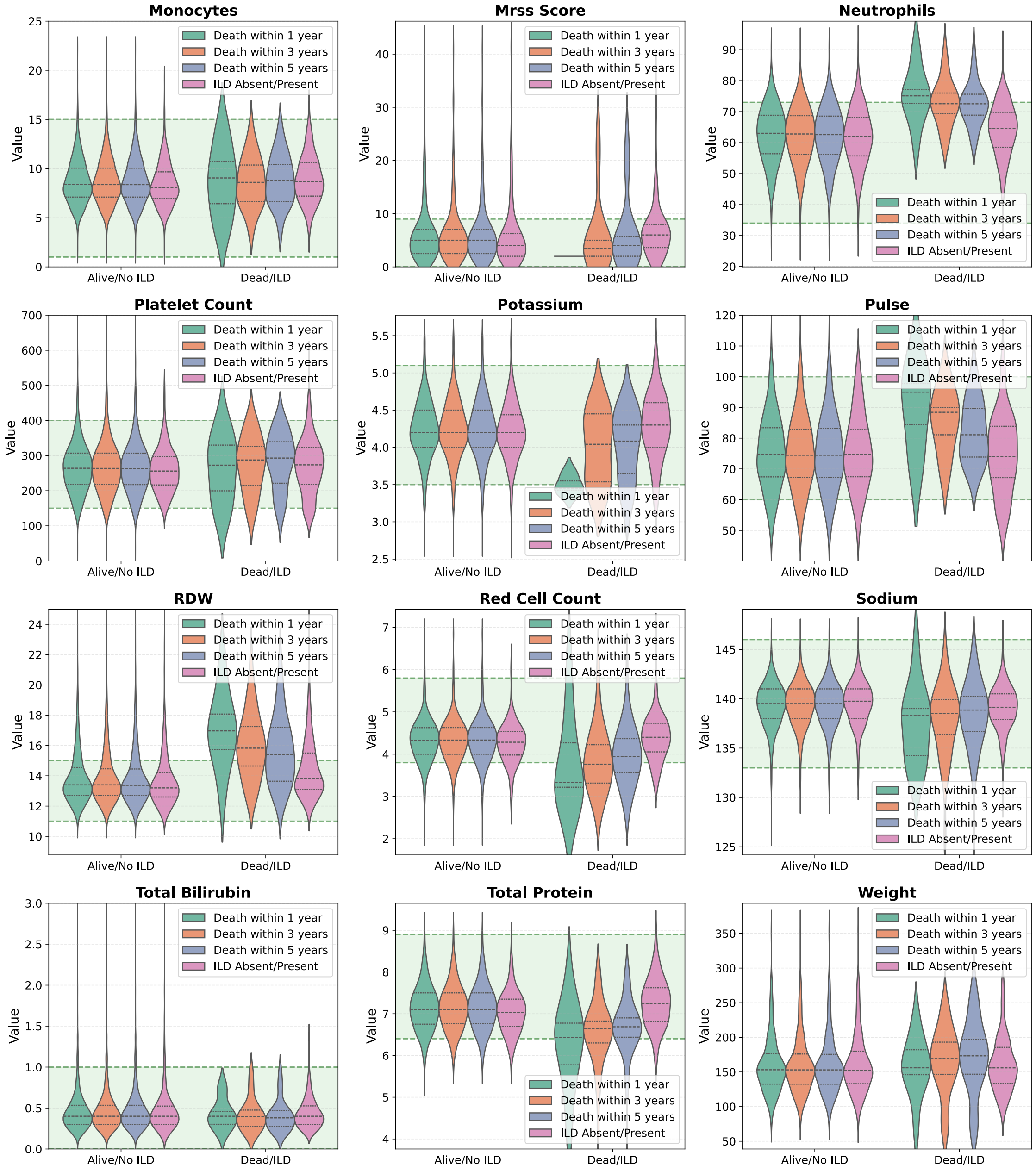

Figure S15: **(Panel D)** Continued from Figure S15C.

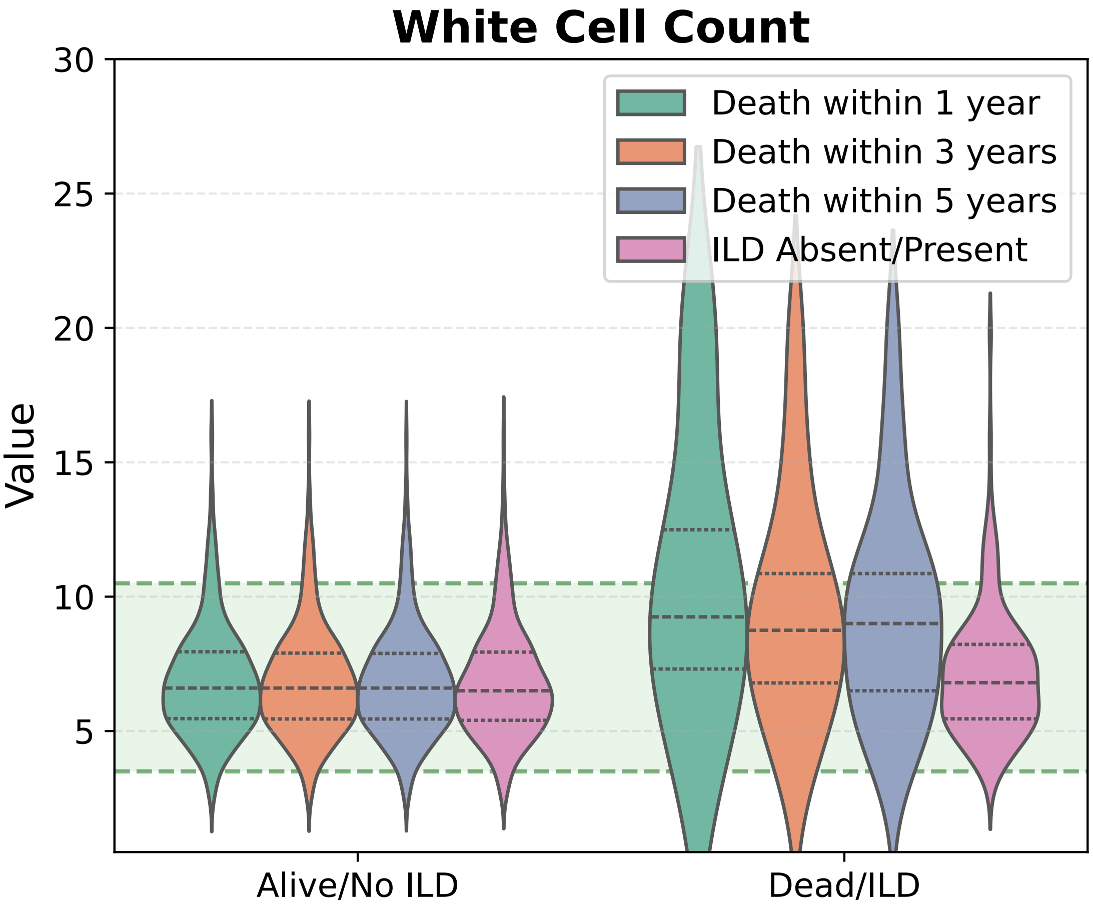

Figure S15: **(Panel E)** Continued from Figure S15D.
